# Evaluating metagenomics and targeted approaches for diagnosis and surveillance of viruses

**DOI:** 10.1101/2024.03.28.24304905

**Authors:** Sarah Buddle, Leysa Forrest, Naomi Akinsuyi, Luz Marina Martin Bernal, Tony Brooks, Cristina Venturini, Charles Miller, Julianne R Brown, Nathaniel Storey, Laura Atkinson, Timothy Best, Sunando Roy, Sian Goldsworthy, Sergi Castellano, Peter Simmonds, Heli Harvala, Tanya Golubchik, Rachel Williams, Judith Breuer, Sofia Morfopoulou, Oscar Enrique Torres Montaguth

## Abstract

**Background:** Metagenomics is a powerful approach for the detection of unknown and novel pathogens. Workflows based on Illumina short-read sequencing are becoming established in diagnostic laboratories. However, barriers to broader take-up include the need for high sequencing depths, long turnaround times, and limited sensitivity. Newer metagenomics protocols based on Oxford Nanopore Technologies (ONT) sequencing allow acquisition and analysis of data in real time, potentially reducing the need for high-volume sequencing and enabling point-of-care testing. Furthermore, targeted approaches that selectively amplify known pathogens could improve sensitivity.

**Methods:** We evaluated detection of viruses with readily available untargeted metagenomic workflows using Illumina and ONT, and an Illumina-based enrichment approach using the Twist Biosciences Comprehensive Viral Research Panel (VRP), which targets 3153 viruses. We tested samples consisting of a dilution series of a six-virus mock community in a human DNA/RNA background, designed to resemble clinical specimens with low microbial abundance and high host content. Protocols were designed to retain the host transcriptome, since this could help confirm the absence of infectious agents. We further compared the performance of commonly used taxonomic classifiers.

**Results:** Capture with the Twist VRP increased sensitivity by at least 10-100-fold over untargeted sequencing, making it suitable for the detection of low viral loads (60 genome copies per ml (gc/ml)), but additional methods may be needed in a diagnostic setting to detect untargeted organisms. While untargeted ONT had good sensitivity at high viral loads (60,000 gc/ml), at lower viral loads (600-6,000 gc/ml), longer and more costly sequencing runs would be required to achieve sensitivities comparable to the untargeted Illumina protocol. Untargeted ONT provided better specificity than untargeted Illumina sequencing. However, the application of robust thresholds standardized results between taxonomic classifiers. Host gene expression analysis is optimal with untargeted Illumina sequencing but possible with both the VRP and ONT.

**Conclusions:** Metagenomics has the potential to become standard-of-care in diagnostics and is a powerful tool for the discovery of emerging pathogens. Untargeted Illumina and ONT metagenomics and capture with the Twist VRP have different advantages with respect to sensitivity, specificity, turnaround time and cost, and the optimal method will depend on the clinical context.

## Background

Metagenomics, the sequencing of all genomic material within a sample, is a demonstrably powerful approach for detection of novel or unknown pathogens. Most notably, metagenomic sequencing identified the SARS-CoV-2 virus within four weeks of the first reported patient being hospitalized (1). The unselective and comprehensive approach makes metagenomics attractive as a diagnostic tool. A single test that can identify any pathogen, including those that are unexpected and novel, holds much interest for clinical and public health laboratories. Since 2008, short read metagenomics has been trialed by many groups to identify causes of fever and central nervous system diseases, including encephalitis, particularly in undiagnosed immunocompromised patients or outbreaks of unknown aetiology (2–11). With the recent advent of rapid methods, such as sequencing with Oxford Nanopore Technologies (ONT), metagenomic approaches have been proposed as suitable for rapid detection of unexpected pathogens and antimicrobial resistance in respiratory samples from patients with complex pneumonias receiving intensive care treatment (12–18). As an augmentation to metagenomics, oligonucleotide panels that enrich for large numbers of pathogens, while potentially reducing the possibilities for detection of an unknown pathogen, have been reported to improve the sensitivity and speed with which known pathogen genomes are detected, making them potentially valuable for infection diagnosis and screening (19–24).

Comprehensive evaluation of these pipelines is vital for their wider uptake in clinical laboratories. A major problem for the routine use of metagenomics in the diagnosis of infection has been the dilemma of distinguishing true and contaminating infectious agents. This is particularly challenging where deep sequencing of material with normally low microbial abundance is required to exclude infection, for example in the differential diagnosis of encephalitis (11). In such cases, absence of a pathogen is as important as its presence, allowing clinical teams to focus on immunomodulatory approaches that could be detrimental if infection is present. The plethora of bioinformatic tools available for interpretation of results and the lack of standardization poses further uncertainties for diagnostic labs and complicates comparison of metagenomic results, particularly if generated by different protocols (25,26).

Several benchmarking studies have been performed comparing long and short read platforms (27–34) and associated bioinformatics methods (25,26,35–40) for bacterial and fungal detection. However, failure to detect viral infections may hinder the utility of metagenomic methods, particularly for the diagnosis of infections in the central nervous system and in patients with compromised immune systems, in whom serious viral infections are a major cause of morbidity and mortality (41–43). Sensitive detection of viruses is also required in other situations, including for example, screening of blood and organs for transplantation (44) and reliably detecting pathogens of high consequence in returning travelers (45,46). A recent study compared viral detection in simulated low biomass samples (e.g. respiratory swabs and CSF) using Illumina, ONT and targeted methods across multiple centres (47). However, high biomass samples, such as blood and tissue, present different technical challenges due to the high levels of host genetic material and may require different metagenomics protocols.

Many metagenomic methods advocate depletion of host nucleic acid to improve sensitivity, especially where microbial abundance is low (48,49). However, depletion significatively reduces host transcriptomic information, which can, when combined with pathogen metagenomics, improve accuracy of diagnosis and provide important insights that inform patient management (27,50–53). Human transcriptomic analysis can identify immune pathways upregulated in the host and can help distinguish between viral, bacterial, and non-infectious causes of disease, which is particularly important when no pathogens are detected through metagenomics (27,50–53). Nucleic acid depletion methods also reduce sensitivity to microbes without cell walls, increase contamination due to additional reagents and reduce sensitivity for detection of cell-free DNA and RNA (54,55).

To provide a pragmatic assessment of utility for routine diagnostic viral metagenomics in samples expected to have low microbial abundance, including blood and tissue, we evaluated three commonly used metagenomic platforms and eight off-the-shelf bioinformatic methods. We established the sensitivity and limits of detection of all methods on a panel of known viral sequences. In addition, we demonstrated modifications that can be used to standardize the outputs of bioinformatic tools and minimize the presence of low-level contaminating microorganisms. This will better enable comparison between different platforms and bioinformatic tools and increase confidence in reporting results. Since combined host-pathogen genomic analysis is increasingly likely to contribute to optimum patient management, we also evaluated how well the methods preserve RNA sequences from the host transcriptome. Our goal is to provide guidance on the capabilities and drawbacks of each, for routine diagnostic use and public health screening.

## Results

### Sensitivity and limit of detection

We tested simulated post-extraction clinical samples where the input viral composition is known, consisting of a mock community of genomic DNA/RNA from six viruses, two DNA (human mastadenovirus F and human betaherpesvirus 5) and four RNA (mammalian orthoreovirus, human orthopneumovirus, influenza B virus and Zika Virus), at four different concentrations in a constant human DNA and RNA background (**Figure 1**). The same input was used for all metagenomics approaches: the untargeted Illumina and ONT protocols, and the capture probe enrichment with the Twist Comprehensive Viral Research Panel (VRP) followed by Illumina sequencing (**Figure 1**). At least two replicates were tested for each technology-concentration pair. We obtained 38.2-81.2 and 38.8-66.9 million reads per sample for untargeted Illumina sequencing and Illumina following the Twist VRP respectively, corresponding to 5.7-12.2 Gb and 5.8-10.0 Gb respectively (**Supplementary Table 1**). For the untargeted ONT sequencing, we obtained 5.1-12.3 Gb per sample. To improve comparability between methods, we randomly subsampled 5 Gb from each sample across the platforms for analysis.

**Figure 1:**
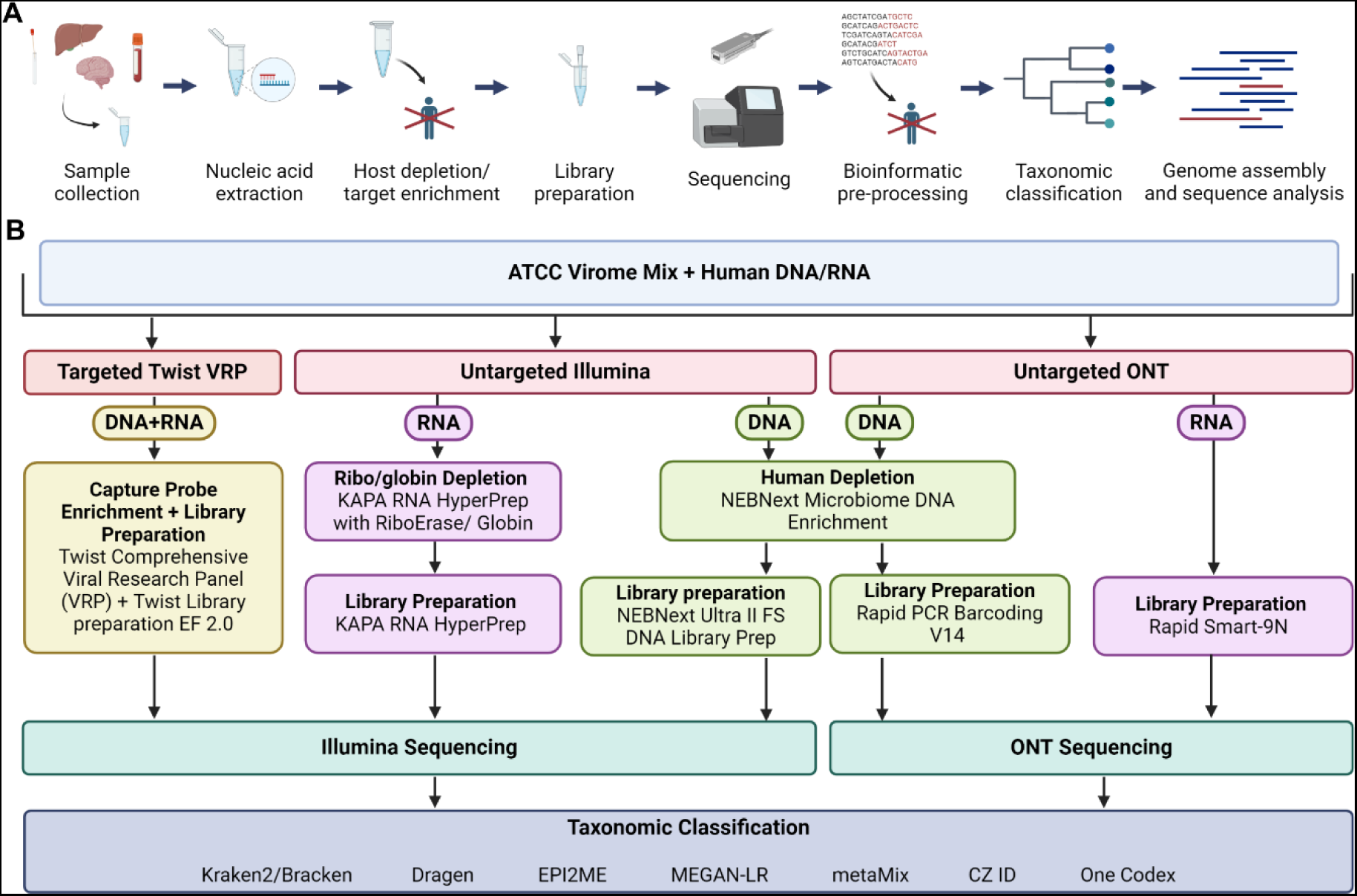
Metagenomic sequencing and experimental outline. **A** Overview of metagenomic processing pipeline. **B** Flow chart summarizing experimental design, which involves inputting mock and clinical samples into three metagenomic workflows: Illumina DNA and RNA seq using NEBNext and KAPA kits respectively, ONT DNA and RNA seq using the Rapid PCR barcoding kit and the Rapid Smart-9N method respectively, and finally the targeted DNA- and RNA-based Twist viral research panel, sequenced on the Illumina platform. The resulting data was analyzed using different taxonomic classifiers. Produced with biorender.com.

The Twist VRP was the most sensitive method, as it was the only platform to detect all the expected viruses at 60 genome copies per ml (gc/ml), with coverage over 98.8% for all viruses at 60,000 gc/ml and ranging from 3.7-23.0% at 60 gc/ml (**Figure 2A**). ONT was less sensitive than Illumina, detecting in at least one of the replicates all six viruses at 60,000 gc/ml, four of six viruses (human betaherpesvirus 5, human mastadenovirus F, orthopneumovirus and Zika virus) at 6000 gc/ml but only two viruses, one double-stranded (ds) DNA (human betaherpesvirus 5) and the other dsRNA virus orthoreovirus at 600 gc/ml and none at 60 gc/ml. The detection of the dsRNA virus orthoreovirus at 600 gc/ml despite not being detected at 6000 gc/ml, represents only four reads in one of the replicates, with no reads detected in the other replicate, likely reflecting stochastic variation. In contrast, untargeted Illumina detected all six viruses at 60,000 and 6,000 gc/ml, five at 600 gc/ml (all apart from human mastadenovirus F) and one at 60 gc/ml (human betaherpesvirus 5) (Error! Reference source not found. **2A**). At levels close to the limits of detection, there was sometimes variation between the technical replicates in their ability to detect the viruses (**Figure 2**). One additional DNA virus (human mastadenovirus F) and one additional RNA virus in one of the repeats (human orthopneumovirus) were detected by Illumina sequencing at 600 gc/ml when additional sequence data was available beyond 5 Gb (9.6 and 10.7 Gb for DNA and 11.1 Gb for RNA) (**Supplementary Table 1**). Other than this, no additional viruses were detected in the full datasets before subsampling.

**Figure 2:**
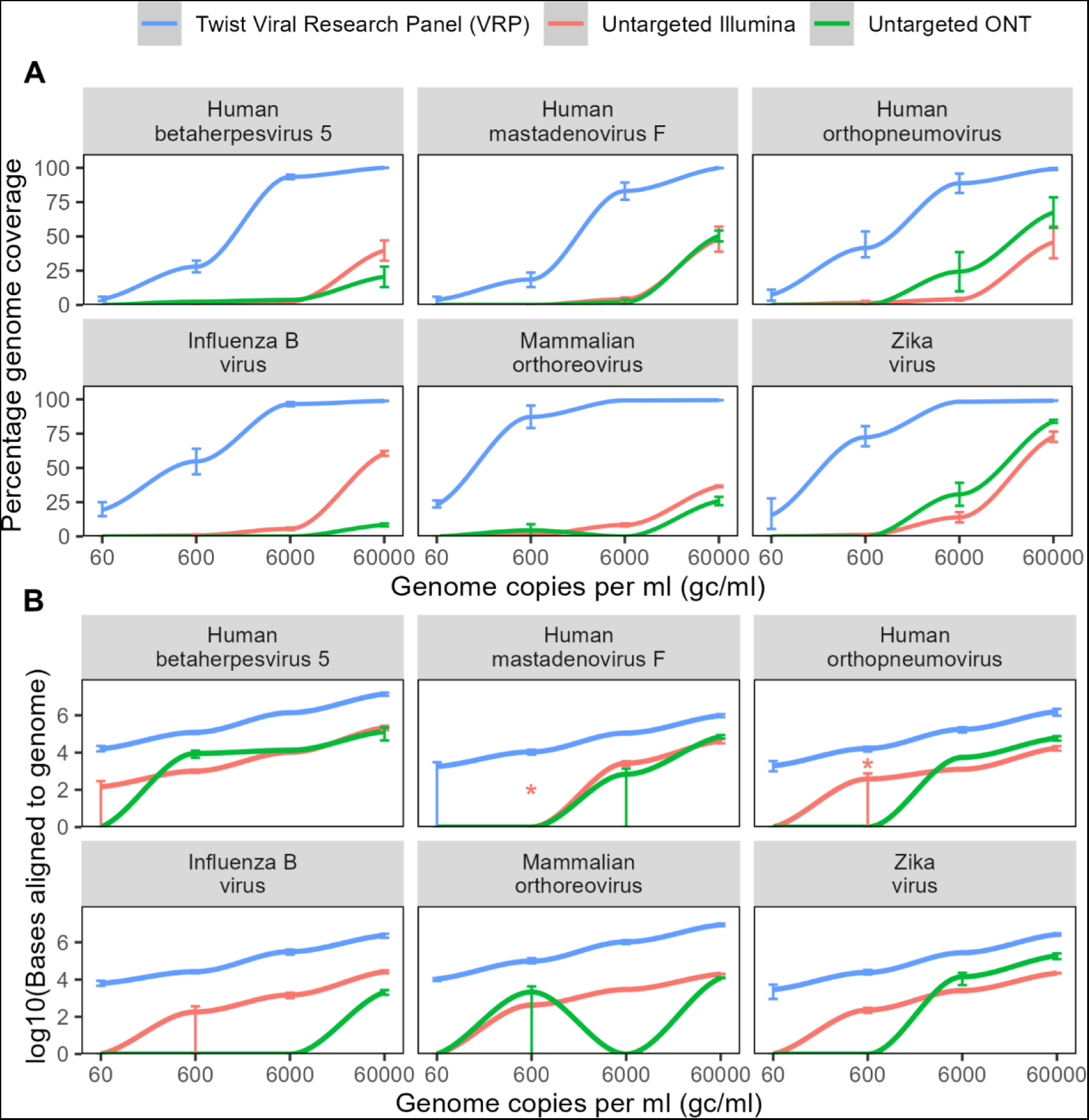
Sensitivity in mock clinical samples. Coverage and base pairs aligned to the six expected viral species in mock samples, by untargeted Illumina and ONT sequencing and capture probe enrichment with the Twist Biosciences Comprehensive Viral Research Panel followed by Illumina sequencing. **A** Percentage genome coverage at depth 1x of species in mock community. **B** log10(bases) aligning to reference genome. Samples where a virus was detected in the full dataset but not the subsampled dataset are indicated with a *. Genome copy numbers refer to an average across the viral species – see **Supplementary Table 3**. Each point shows the mean of at least two technical replicates – error bars show the range. PCR duplicate reads removed.

At 60,000 gc/ml, assigned bases ranged from 17,527-217,630 of 5 Gb for Illumina and 2110-134,026 of 5 Gb for ONT (**Figure 2B**). Both ONT and Illumina untargeted sequencing provided incomplete coverage at all concentrations tested, ranging from 8.4 to 83.9% at 60,000 gc/ml, 0-30.7% at 6000 gc/ml, and 0-8.9% at 600 gc/ml (**Figure 2A**). Viruses with longer genomes were detected in with greater read numbers, however normalizing for genome length gave similar abundance estimates for each virus, where viral loads were high enough for consistent detection (**Supplementary** Figure 1). As expected, all technologies displayed levels of PCR duplication ranging from 0-16%, with the Twist VRP showing the highest rates (**Supplementary** Figure 2A). Including PCR duplicates makes no difference to the conclusions regarding sensitivity (**Supplementary** Figure 2B).

We also tested the sensitivity of a range of taxonomic classifiers. The classifiers tested and reasons for inclusion are outlined in **Table 1**. Where no thresholds were applied, all the classifiers had similar sensitivity, although there was some variation in ability to detect viruses at 60-6000 gc/ml for untargeted Illumina sequencing and at 60,000 gc/ml for untargeted ONT sequencing, with Kraken2, Dragen, metaMix-fast and CZ ID being the most sensitive at these viral loads (**Figure 3A**). MetaMix and MEGAN-LR failed to identify influenza B virus and mammalian orthoreovirus respectively with ONT sequencing at 60,000 gc/ml; both RNA viruses for which fewer than 10 reads were detected by the aligner minimap2. Of the other classifiers, One Codex had substantially lower sensitivity for the Twist VRP data compared to other classifiers, all of which identified almost all the viruses at all concentrations tested (**Figure 3A**). This may be because the program only reports organisms that reach a set of predetermined abundance thresholds (56), which may not be reached at low viral loads, whilst the other classifiers do not by default use such thresholds. Where viruses were detected, the classifiers provided broadly similar estimates of reads per million, ranging, for example, from 30.3-73.6, 31.0-39.2 and 3695-8164 RPM, for human betaherpesvirus 5 for Illumina, ONT and the Twist VRP respectively (nucleotide-based classifiers only) (**Supplementary** Figure 3).

**Figure 3:**
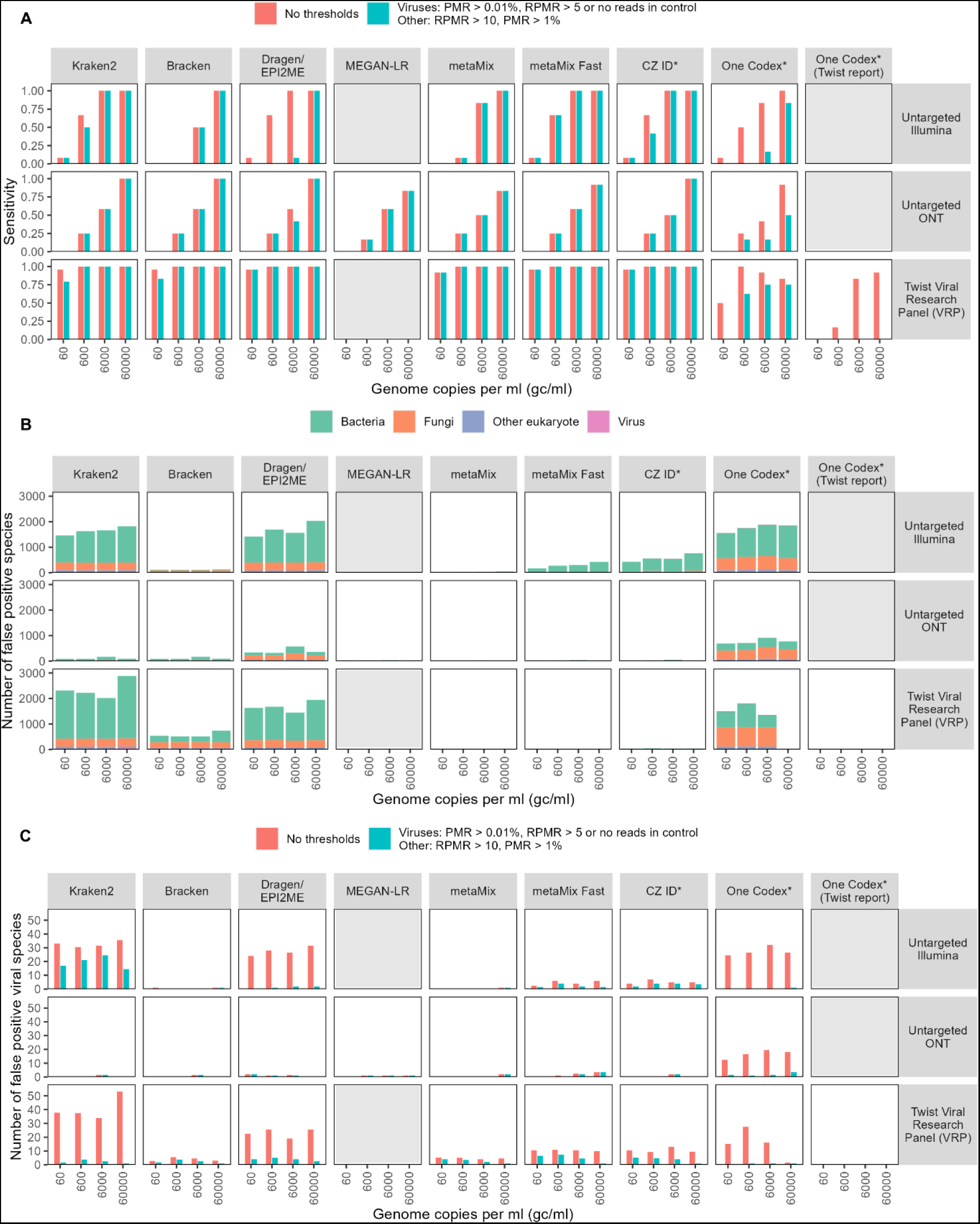
Sensitivity and specificity of taxonomic classifiers. **A** Sensitivity to the species in the mock community before and after the application of thresholds, for seven different taxonomic classifiers, by untargeted Illumina and ONT sequencing and capture probe enrichment with the Twist Biosciences Comprehensive Viral Research Panel followed by Illumina sequencing. MEGAN-LR and the One Codex Twist report are only designed for ONT and Twist sequencing respectively so were only run for these platforms. **B,C** Number of false positive species, defined as a species that is classified as positive but not present in the mock community **B** False positive species from the raw output of the taxonomic classifiers with no thresholds applied. **C** Comparison of the numbers of viral positive species identified before and after the application of thresholds. Genome copy numbers refer to an average across the viral species – see **Supplementary Table 3**. Each bar shows the mean of at least two technical replicates.

**Table 1:** Taxonomic classifiers.

| Classifier | Method | Reason included | Platform | GUI or CLI | Local or cloud | Database size (based on identical fasta files) | Approximate time taken (hours)* | Reference |
| --- | --- | --- | --- | --- | --- | --- | --- | --- |
| **Kraken2 & Bracken** | Kmer-based, lowest common ancestor | Very widely used | Illumina & ONT | CLI | Local | 124 GB | 1-3 | (58,59) |
| **DRAGEN Metagenomics (Kraken2)** | See Kraken2 | Illumina's platform | Illumina & ONT | CLI & GUI | Cloud | 124 GB | 1.5-3 | (60) |
| **EPI2ME Labs wf-metagenomics (Kraken2 & Bracken)** | See Kraken2 | ONT's platform | ONT | CLI & GUI | Local. Cloud in development | 124 GB | 0.5-2 | (61) |
| **MEGAN-LR** | Lowest common ancestor | Good performance in benchmarking study(39) | Illumina & ONT | CLI required for preprocessing. GUI (free), CLI (paid for short reads) | Local | 327 GB (Minimap2)<br>88 GB (DIAMOND) | 5-8 | (62) |
| **metaMix** | Bayesian mixture models | Good performance in benchmarking study, used clinically(36) | Illumina & ONT | CLI | Local | 148 GB (BLAST)<br>88 GB (DIAMOND) | 5-12+ | (63) |
| **CZ ID** | Alignment and assembly | Free, cloud-based platform | Illumina & ONT | CLI & GUI | Cloud | NA – inbuilt online database | 0.5-2 | (64) |
| **One Codex** | Kmer-based | Recommended platform for use with Twist VRP | Illumina & ONT | CLI & GUI | Cloud | NA – inbuilt online database | ~0.5-2 | (65) |
| **Kaiju** | Local alignment based | Widely used protein classifier | Illumina & ONT | CLI | Local | 101 GB | 1-3 | (57) |

\* Shows approximate range only – exact time taken depends on sample complexity, number of samples processed and computational resources, and exact timings were not available for all classifiers, e.g One Codex. Based time taken to process single samples from our dataset, including bioinformatic preprocessing.

### Specificity and false positive rates

High precision and low false positive rates are as important as sensitivity in a clinical diagnostic setting, and rational approaches to identifying and reporting contaminants, particularly by non-specialist bioinformaticians, are needed. To evaluate their performance, we compared the number of false positive species identified by a range of commonly used taxonomic classifiers for the mock samples (**Table 1**). A false positive is defined as any species not present in the mock community. All the classifiers assigned similar numbers of reads to the species in the mock community, except for One Codex, which had lower sensitivity for the Twist VRP data than the other classifiers. (**Supplementary** Figure 3). However, when no additional thresholds were applied, there was a large variability between the classifiers in terms of the number of species identified by Illumina sequencing (**Figure 1B)**. Most of the false positive species were fungi or bacteria. Kraken2 Illumina’s Dragen Metagenomics Pipeline (which is based on Kraken2) and One Codex, all use kmer methodologies and identified over 1500 false positive species for the untargeted Illumina sequencing. The discrepancy between the number of false positives identified for the Twist VRP data by One Codex at different concentrations may be caused by greater availability of data for the classifier to distinguish between true and false positives at higher read depths (56). By contrast, metaMix and Bracken, which both use Bayesian methods, identified only one false positive viral species at 60,000 gc/ml, (**Figure 3A&C**). However, both these classifiers were less sensitive at lower genome copy numbers than classifiers such as Kraken2 and CZ ID. In contrast to Illumina, few false positive species, especially viruses, were identified with ONT sequencing. Thus, for ONT the application of thresholds beyond a basic comparison to the negative control may not be required.

To reduce the number of false positive species identified for Illumina sequencing, we imposed more stringent thresholds. Completely disregarding all species with any reads in the negative control may result in a reduction in sensitivity, particularly when there is low-level cross-contamination from high viral load samples into the control. We found that using a combination of reads per million ratio between sample and the corresponding negative control and proportion of microbial reads resulted in optimum sensitivity (91.7%) and specificity (77.4%), which may be useful for classifiers such as Kraken2 and One Codex which require additional thresholds (**Figure 1A&C**). In contrast, ONT sequencing and classifiers such as metaMix have few false positive reads and can be used with only a comparison to the negative control. More details of the derivation of our thresholds can be found in the supplementary information. Use of protein-based classifiers, including Kaiju (57) and the protein modes of MEGAN-LR, metaMix and CZ ID, did not improve the sensitivity or specificity of classification (**Supplementary** Figure 4). The false positive viral species that remained after the application of thresholds were mainly viruses that do not infect mammals or birds, making them unlikely to be clinically relevant (**Supplementary** Figure 5). The remaining false positive viruses were mainly Anelloviridae (often Torque Teno viruses), and viruses that were related to those in the mock community, such as other herpes or adenoviruses. The Anelloviridae, which are very commonly found in human samples, were mostly not found in the controls and are possibly a result of low-level contamination.

### Host transcriptomic analysis

Several studies highlight the power of host transcriptomics methods for distinguishing bacterial, viral, and non-infectious causes of illness (27,50–53), although none are being used diagnostically at present. When metagenomics does not identify any pathogens, such analysis could help distinguish between a non-infectious cause of disease and a lack of sensitivity of the metagenomics protocol. Since Illumina RNA sequencing has been extensively used and validated for transcriptomic studies, we compared the estimates of human gene expression provided by the ONT and Twist VRP platforms to those from Illumina. Although the Twist VRP only enriches for viruses, it retains the background, meaning that this analysis remains possible. The number of reads assigned to each human protein-coding gene were positively correlated between Illumina and the other two technologies (correlation coefficients, Spearman’s rho, 0.694 and 0.709 for ONT and the Twist VRP respectively) (**Figure 4A-C**). Due to the combined DNA and RNA protocol used with the Twist VRP, there were a large number of human genes that were identified as highly expressed by the panel but not untargeted Illumina (**Figure 4B**). We therefore repeated the analysis, focusing only on reads that mapped across exon-exon junctions, termed henceforth “spliced reads”, which are likely to represent mRNA, resulting in a better agreement between the Illumina and the Twist VRP results (**Figure 4D**).

**Figure 4:**
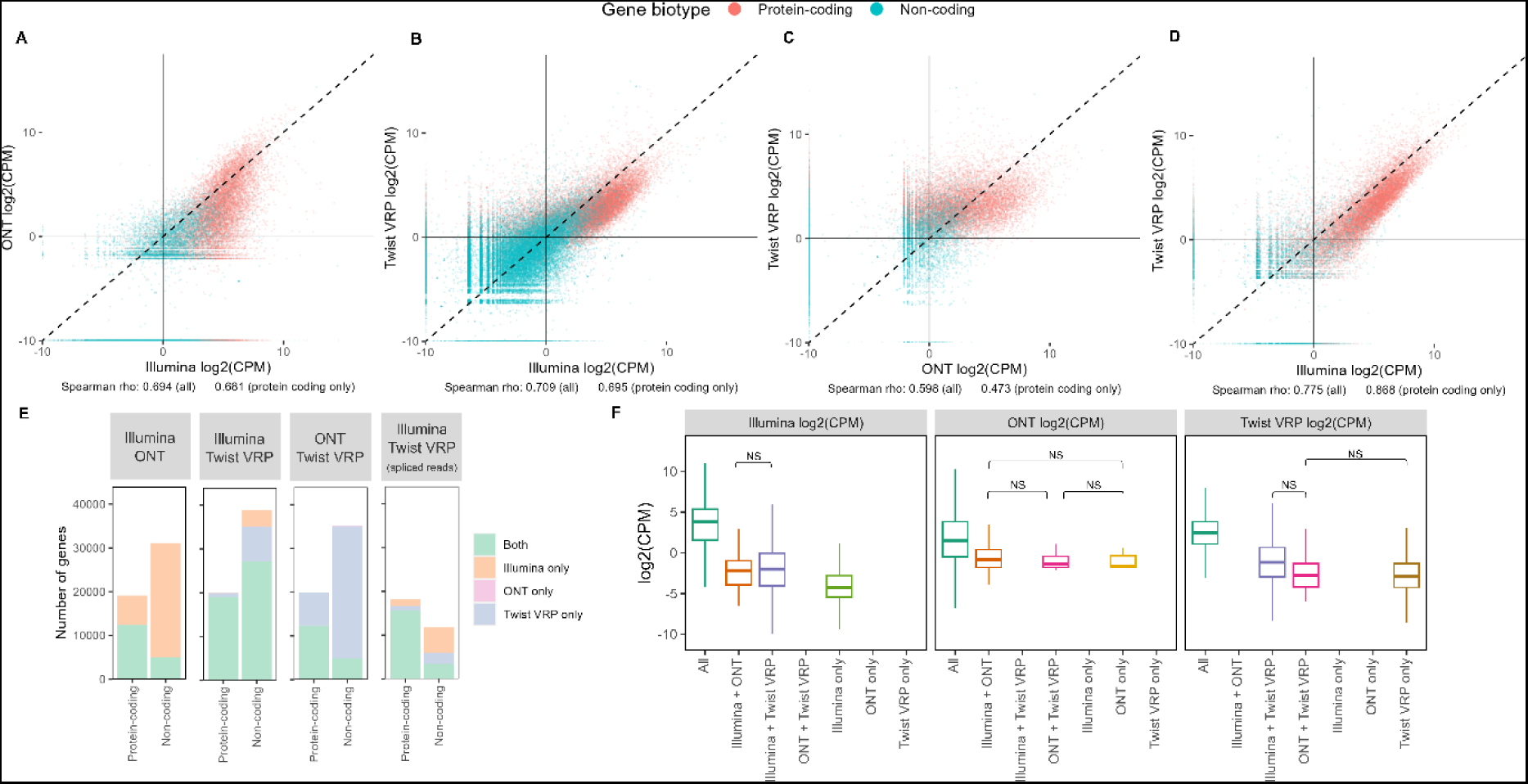
Host transcriptomic analysis. **A-D** Read counts per million assigned to each gene in the human genome by untargeted Illumina, untargeted ONT and targeted Illumina sequencing using the Twist Viral Research Panel. Each point represents a gene**. A-C** raw reads; **D** only reads that map across splice junctions. **E** number of genes identified by each pair of technologies. **F** counts per million of reads by platform. Each panel shows the log2(CPM) as estimated by a different technology. Outliers not shown. All comparisons are statistically significant (p < 0.01) with a pairwise Wilcox test other than those indicated.

While most protein-coding genes were identified by all the technologies (**Figure 4E**), there was still a substantial minority that were not identified by ONT (**Figure 4E**). Use of spliced reads for untargeted Illumina and Twist VRP, only resulted in a small drop in the number of protein-coding genes identified, and a larger drop in the non-coding transcripts (**Figure 4E**). Genes that were identified by all technologies were significantly more highly expressed (**Figure 4F**) suggesting that low-expressed genes may be less reliably identified by all technologies, particularly ONT.

### Turnaround time and cost

Costs and turnaround times from sample to results affect the adoption of metagenomics for routine diagnostics. ONT provides the quickest library preparation method, at just over 5 hours for both DNA and RNA protocols (**Figure 5A**). Targeted sequencing with the Twist VRP requires overnight hybridization and is the slowest protocol (**Figure 5A**). The Twist VRP protocol was the cheapest based on 23 samples (+ negative control) and a sequencing depth of 5 Gb, while the untargeted Illumina sequencing was the most expensive (**Figure 5B, Supplementary Table 2**).

**Figure 5:**
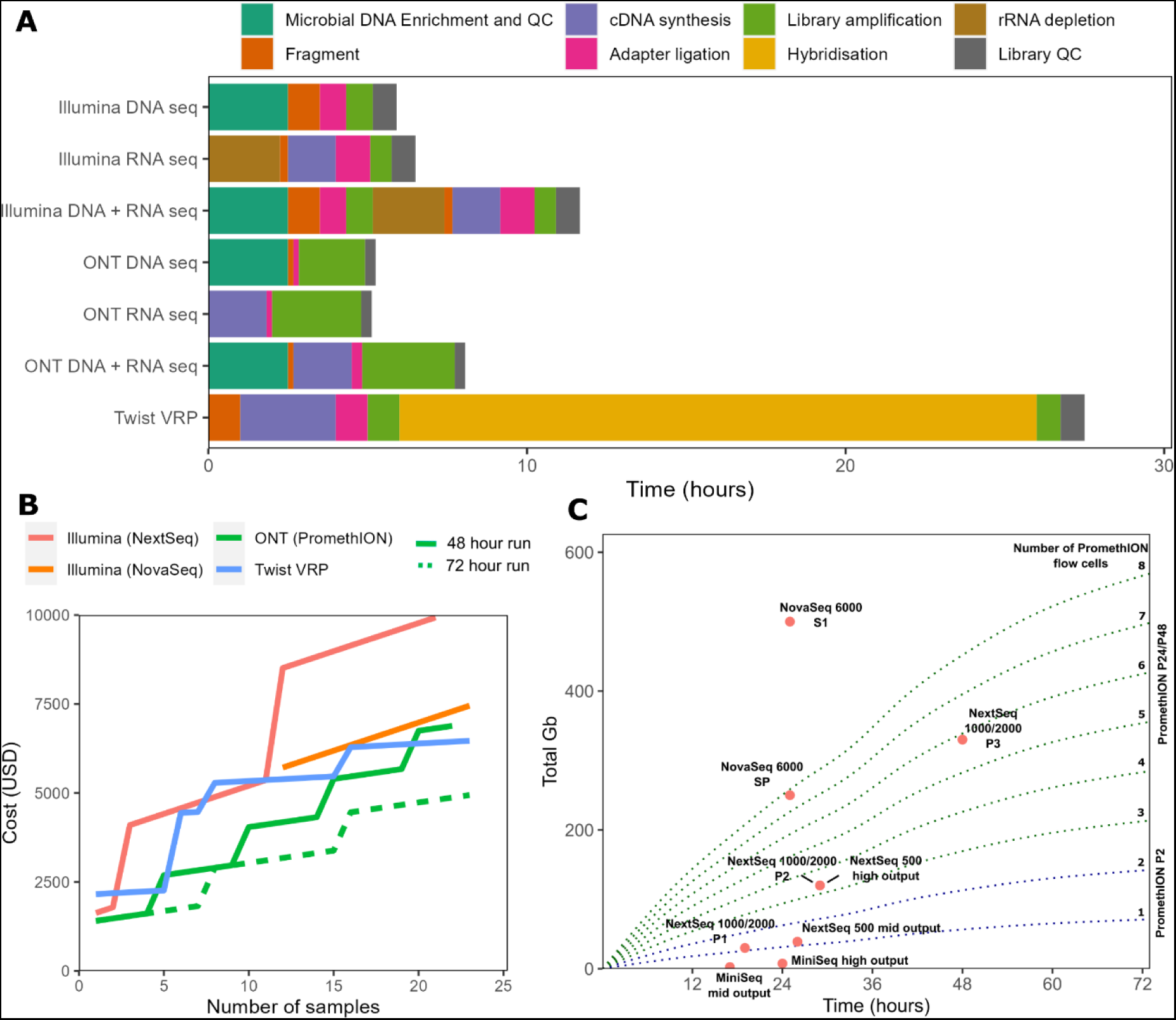
Turnaround times and output data volumes. **A** Time taken for library preparation for the different protocols tested. The Twist panel uses a combined DNA and RNA-Seq protocol. The DNA+RNA bars for the untargeted sequencing indicate the time taken if both protocols are performed by a single operator. **B** Total cost (including library preparation) to sequence number of samples indicated plus single negative control, to a depth of 5GB. ONT costs are shown with 48- and 72-hour maximum run times per flow cell. **C** Volume of data output by time for a range of Illumina sequencing kits and ONT sequencing with PromethION flow cells. The Illumina kits produce a set amount of data after the sequencing run is complete – this is shown by pink dots. In ONT sequencing, data is output continuously and the run can be stopped at any time, until the flow cell becomes degraded. PromethION data (green/blue dotted lines) shows the average of our RNA and DNA-Seq runs, passed reads only. Data outputs for Illumina were obtained from the product specification data as of April 2024.

However, directly comparing the costs of each protocol to obtain at least 5 Gb of sequence data does not account for differences in their sensitivity. Since the Twist VRP approach is at least 10-100x more sensitive than untargeted Illumina and 100-1000x more than untargeted ONT (**Figure 2**), increases of orders of magnitude in sequencing depth would be required to bring the sensitivity of the untargeted protocols in line with that of the Twist VRP. Even up to twice as much sequence data (10.6 Gb and 11.3 Gb at 60 gc/ml) did not increase the sensitivity of untargeted Illumina and ONT respectively to anything near to the Twist VRP. For untargeted Illumina, greater sequencing depth also amplifies the detection of contaminants, making interpretation more difficult. Achieving increased sensitivity using ONT sequencing would require long sequencing runs and a reduction in the number of samples sequenced per flow cell, significantly increasing costs and turnaround times. This means that targeted metagenomics methods such as Twist VRP are by far the quickest and most cost-effective of the protocols for detection of low viral loads (60-600 gc/ml). Similarly, since untargeted Illumina is more sensitive than this untargeted ONT protocol, it will be quicker and cheaper to reach the sequencing depths required to detect intermediate viral loads (600-6000 gc/ml) using Illumina.

Sequencing costs and turnaround times will also be influenced by the number of samples. For fewer than six samples, including controls, ONT is the cheapest and fastest alternative where microbial load is likely to be high and genomic sequences are achievable with lower sequencing depth, for example 5 Gb of data, per sample (**Figure 5A&B**). ONT also provides access to the sequencing data in real time, allowing preliminary analysis of the results before the run is completed, which can be advantageous for samples with high viral loads. However, if more samples are processed in parallel or a higher sequencing depth is required to improve sensitivity to a level comparable with untargeted Illumina, longer sequencing runs will be needed (**Figure 5B&C).** When the total volume of data required per run is higher than around 30 Gb, it may be faster to use Illumina sequencing (**Figure 5C**). However, it remains cheaper to use ONT with 23 sample runs (24 including negative control, 120 Gb) (**Figure 5B**). Because of the Twist VRP’s improved sensitivity, lower sequencing depths are required per sample, allowing the use of smaller Illumina sequencers and cheaper kits with shorter sequencing times (**Figure 5C**). However, for fewer samples the Twist VRP method may be much more expensive, since the kit optimal cost per sample is based on the pooling of 7 (8 including negative control) samples per hybridization (**Figure 5B, Supplementary Table 2)**.

## Discussion

The use of metagenomics and allied targeted methods for routine diagnostics and clinical management are now priorities for laboratories in many countries. At least two commercial solutions are already available, in both cases using Illumina platforms for untargeted sequencing of cell-free DNA in blood, to identify causes of sepsis (67,68). However, these approaches may not be suitable for the detection of cell-associated pathogens, notably viruses, and data on limits of detection for viruses is absent. Untargeted Illumina sequencing is also in routine use in a handful of labs for the management of patients with fever of unknown origin, encephalitis, meningitis, and sepsis (10,11,69). Most recently, routine diagnostic ONT metagenomic sequencing of respiratory samples has been proposed for improved management of critically ill patients with pneumonia (12,13). In each case the metagenomic set-ups are multi-step workflows where each stage, from sample collection to computational data analysis, significantly affects the outcome of the test (26,28,70) (**Figure 1A**). Sensitivity, specificity, reproducibility, turnaround time and cost are critical considerations before implementation in a clinical laboratory. However, with limited standardization across workflows, few head-to-head comparisons and significant, if underreported, drawbacks to most of the existing pipelines, choosing and implementing a metagenomics workflow remains complicated and uncertain for most.

In this study we have focused on detection of viruses, which are particularly important causes of morbidity and mortality in immunocompromised patients (41–43). Sensitive detection of viral infections is also necessary where metagenomics is being considered for screening of biological therapies such as blood and organ donations (44) and for detection of pathogens of high consequence, for example in returning travelers (45,46). Detection of viruses also has implications for antimicrobial stewardship and with increasing antiviral agents available, the appropriate stratification of patient management. Several studies have previously compared Illumina and ONT-based metagenomics of bacterial and fungal mock communities (29,30,32), simulated bacterial datasets (32,33,38) and clinical samples (18,28,71,72). Some work on viral detection from clinical samples (27,73–77) or mock communities resembling environmental samples (34) has also been reported. While the sensitivity of both platforms, where compared, has been found to be similar for bacterial detection (27,29,30,38,73,78), few have compared detection of RNA viruses. Recently, a multicenter study benchmarking 11 clinical metagenomic workflows using a panel of simulated low biomass samples, including CSF and nasopharyngeal swabs, tested different viral loads and showed that only a minority of protocols, including a Twist VRP approach, were able to detect viruses at CT values of over 35 (47). However, to our knowledge, no studies have systematically tested different viral loads, established limits of detection or specificity for viral detection and evaluated the quality of host transcriptomics information in samples with high human background.

Here we show that untargeted Illumina and ONT metagenomics, and targeted Illumina sequencing with the Twist VRP, detect high viral loads (60,000 gc/ml) with good sensitivity and reproducibility. Untargeted Illumina sequencing appears better able than ONT to detect viruses at lower genome copy numbers, with the former finding all six viruses at 6000 and five at 600 gc/ml, while the latter detected only four and two of the six viruses respectively, with only untargeted Illumina finding a single virus at 60 gc/ml (**Figure 2**). Notably ONT detected only two of the four RNA viruses at 6000 gc/ml, one of the four at 600 gc/ml and none at 60 gc/ml. This may be because depletion of ribosomal RNA (rRNA) before performing Rapid-SMART-9N (79), which is known to improve Illumina detection of RNA viruses, resulted in levels of RNA input that are too low for adequate ONT library preparation. In order to overcome this, adapting the current workflow to include cDNA synthesis kits compatible with ultra-low input RNA should be considered, which could improve the sensitivity of ONT, particularly for single stranded (ss) RNA viruses. Combining ONT with differential lysis methods, which remove host and non-encapsulated nucleic acids, can improve sensitivity (80,81) detection of bacteria and fungi, but this step may reduce sensitivity for certain microbes and reduce the ability to detect cell-free DNA and RNA, including viral nucleic acid (54,55). Furthermore, with increasing moves to combine host gene expression with microbial detection to improve infection-diagnosis rates (27,50–53), methods such as differential lysis, which deplete human nucleic acid may be less attractive.

More sensitive than either untargeted Illumina or ONT, viral enrichment using the commercially available Twist VRP panel was able to detect all six viruses down to levels of 60 gc/ml, a finding in keeping with reports for other commercial capture protocols (19). However, the Twist VRP only includes viral probes and may require the addition and evaluation of probes targeting other pathogens and AMR genes to be useful for routine diagnostic use, since a virus-only panel does not allow syndromic diagnosis of infection. Having a defined panel may also limit the ability to detect novel pathogens. The probes can detect organisms with up to 20% difference to the reference with over 50% coverage (82), but cannot detect more divergent infectious agents, as exemplified by the failure of the Twist VRP to detect the internal control *E. coli* phages Lambda and MS2 (0-32 reads with the Twist VRP versus 1322-1950 reads with untargeted Illumina at 60,000 gc/ml for MS2 phage). However, as demonstrated by the host transcriptomic analysis of the Twist VRP data, non-targeted material is retained by this protocol. This means that it may be possible to detect non-targeted microbial species, including bacteria, fungi, and highly divergent viruses, if their abundance is high enough in relation to the depth of sequencing used. Finally, capture probe-based methods are currently designed only for use with Illumina sequencing. Previously reported attempts to add an enrichment step to improve the sensitivity of ONT sequencing require first generating an Illumina sequencing library before converting this for ONT sequencing through additional library preparation steps (22), making this approach costly and time-consuming.

The propensity for deep sequencing metagenomic methods to detect contaminant species presents a particular challenge when such methods are considered for routine diagnostic use. The numbers of falsely detected species were lowest for ONT sequencing and greatest for untargeted Illumina sequencing and the Twist VRP (**Figure 3B**). The higher precision of ONT is most likely due to longer reads making it easier for taxonomic classifiers to unambiguously assign reads to species. Large numbers of false positive species were identified for untargeted and targeted (Twist VRP) Illumina sequencing. This was particularly pronounced for commonly used classifiers for bacterial data such Kraken2, whose kmer-based approach can result in inaccurate assignment of short reads due to cross-mapping (**Supplementary** Figure 5) (39). Although the results of Kraken2 can be improved by post-processing with Bracken, this approach has a lower sensitivity than classifiers such as metaMix and CZ ID.

By contrast, the use of probabilistic methods that inherently control false positives, such as metaMix, reduced the numbers of false positive species to levels similar to those seen for ONT (**Figure 3C**). By applying thresholds based on a combination of reads per million ratio, which compares species detected in samples and corresponding negative controls, and proportion of microbial reads, we demonstrate that false positive rates can be reduced for all classifiers, thus standardizing outputs from different sequencing methods and classifiers. Our approach differs from those previously applied, where only one of these measures or raw read counts alone were used. Our method highlights the importance of sequencing negative controls, which can help remove contaminants, particularly those present in the reagents. Using this approach, our results suggest capture panels, such as the Twist VRP, provide the best sensitivity and specificity for routine detection of viruses, albeit with the caveats discussed above. Importantly we show that the use of suitable taxonomic classifiers or appropriate thresholds based on comparison with the negative control and the proportion of the total reads assigned to that species overcomes the low specificity that has previously been reported for the Twist VRP when used with its recommended One Codex platform (**Figure 3**) (21).

Host transcriptomic data obtained from untargeted Illumina sequencing has been shown to help distinguish between types of pathogen and infectious and non-infectious causes of disease, which could help to confirm negative or inconclusive results from pathogen identification (50,51). The Twist VRP and ONT show relatively good agreement with the untargeted Illumina protocol’s estimates of human gene expression, although ONT fails to detect some low-abundance genes (**Figure 4**). It is therefore likely that useful transcriptomic information may be obtained from any of the protocols, providing a method that preserves human RNA is selected. The analysis remains possible with the Twist VRP because non-targeted DNA/RNA sequences are retained in an unbiased way, even though the targeted viral sequences are enriched.

Both turnaround times and cost are critical parameters when considering the introduction of new diagnostic methods. Targeted sequencing with the Twist VRP was the only viable method we tested for detection of low viral loads (60 gc/ml), since increasing the depth of untargeted sequencing by the orders of magnitude required to match the sensitivity of the Twist VRP is too expensive and time-consuming to be practical. If an untargeted approach is required, perhaps to test for bacteria and other microbes as well as viruses in a single test, ONT can provide rapid results in cases where sample numbers are low and viral loads are high. In most other circumstances, Illumina is currently the quicker and cheaper way to produce the volumes of data required, particularly as higher volumes of ONT data are required to give the same level of sensitivity. Illumina sequencing may also allow more reliable quantification of human gene expression, making it easier to rule out infection when no pathogens are found.

Our study has several limitations. Since we used commercially available purified nucleic acid standards, we do not compare extraction protocols, which have been shown to have a large impact on the results of metagenomics (83,84). Different approaches have been used to reduce host content in samples in efforts to improve sensitivity. Pre-purification methods like filtration and centrifugation can efficiently remove human cells. However, they can significantly reduce sensitivity for cell-associated viruses (85–87). Alternatively, differential lysis-based methods, which rely on selectively lysing human cells either using mechanical methods such as bead-beating (48) or with saponin (88), have been used to deplete human DNA and RNA prior to ONT sequencing. However, these approaches can lead to biases in organisms detected and reduce detection of cell-free DNA, which may arise from organisms killed by the immune system or antibiotics (54). Additionally, any protocol that removes host material during or before the lysis steps, may lead to reduced sensitivity for integrated and intracellular viruses (54,55). These approaches could also be used prior to Illumina sequencing, although they will prevent host transcriptomic analysis.

Furthermore, we focused only on viruses, while the key advantage of metagenomics is its ability to detect all organisms. Although several studies have shown similar sensitivity to bacteria for Illumina and ONT sequencing on mock communities (29,30), further work is needed to compare commonly used methods such as Illumina sequencing of cell-free DNA and ONT sequencing with differential lysis for detection of bacteria and eukaryotic microbes. We focus only on sterile site samples with high host content such as tissue and whole blood. We expect that different laboratory and bioinformatics methods will also be appropriate for non-sterile sites such as respiratory samples and for samples with low biomass such as plasma and CSF.

## Conclusions

Different metagenomics platforms perform best in terms of sensitivity, specificity, and turnaround times, with no single test currently being optimal in all clinical contexts. Where sensitivity for viral detection is less of a consideration, as might be the case for respiratory samples from severely ill patients with pneumonia, ONT is faster and cheaper. Target capture approaches with Illumina may be preferred for samples with low microbial diversity, where high sensitivity for both DNA and RNA viruses is required to reliably confirm or exclude infection, for example in immunosuppressed patients with fever or encephalitis, blood products and where high consequence pathogens are suspected. Development of rapid, commercially available targeted methods for a wide range of pathogens for both long and short read platforms, using methods that preserve the host transcriptome and also allow rapid untargeted metagenomics where required for pathogen discovery, will bring us closer to a diagnostic test that can detect any pathogen in an actionable timeframe and that could revolutionise clinical microbiology.

## Methods

### Mock clinical samples

Mock samples were prepared to represent high-biomass samples (e.g. blood and tissue) with a clinically relevant spectrum of viral loads ranging from 60 to 60,000 gc/ml. This was achieved by performing serial dilutions of a commercial genetic material mix – the ATCC Virome Nucleic Acid Mix (ATCC, MSA-1008) (**Table 2**) in a background of either human DNA, RNA or a DNA+RNA mix at a concentration of 40 ng/µl. Mock samples were produced in a single large batch to reduce variation and stored as 10 µl single-use aliquots. In parallel, a batch of negative controls containing either DNA, RNA or DNA+RNA without any viral nucleic acids were also generated. DNA and samples were spiked with Lambda phage DNA and RNA samples were spiked with MS2 Bacteriophage RNA. DNA + RNA mixed samples were spiked with both standards. Details of mock sample composition can be found in **Supplementary Table 3.**

**Table 2:** Species composition of ATCC virome virus mix.

| Species | DNA or RNA | Average genome GC content | Genome length (nt) | Envelope |
| --- | --- | --- | --- | --- |
| Human mastadenovirus F | DNA | 51.2 | 34392 | Unenveloped |
| Human herpesvirus 5 (aka cytomegalovirus, CMV) | DNA | 57.1 | 229354 | Enveloped |
| Human orthopneumovirus (aka respiratory syncytial virus, RSV) | - RNA | 33.3 | 15228 | Enveloped |
| Influenza B virus | -RNA | 40.1 | 18527 | Enveloped |
| Mammalian orthoreovirus 3 | +/- RNA | 46.9 | 23416 | Unenveloped |
| Zika virus | + RNA | 50.3 | 10952 | Enveloped |

### Untargeted Illumina sequencing

Untargeted Illumina DNA and RNA metagenomic sequencing of the mock clinical samples was performed as previously described (90). Two technical replicates per mock sample were performed. DNA samples underwent human CpG-methylated DNA depletion using the NEBNext^®^ Microbiome DNA Enrichment Kit (New England Biolabs; E2612L) followed by library preparation using the NEBNext® Ultra™ II FS DNA Library Prep Kit for Illumina (New England Biolabs; E7805L). RNA samples underwent ribosomal RNA (rRNA) depletion followed by library preparation using KAPA RNA HyperPrep kit with RiboErase HMR (Roche; KK8561).

All pre-PCR steps were carried out under an MSC class II cabinet and moved to a post-PCR area following amplification. Libraries were quantified with high sensitivity dSDNA kit (Invitrogen; Q33231) on an Invitrogen Qubit 4 Fluorometer and the average peak sizes for libraries were checked using high sensitivity D1000 screentapes (Agilent 5067-5584) on a Tapestation 4200. Samples were sequenced in equimolar pools using a NextSeq 2000 or a NovaSeq 6000 300 cycle kit (2 x 150 bp) depending on the number of samples processed. A minimum output of 5 Gb per sample was obtained (**Supplementary Table 1**).

### ONT sequencing

ONT sequencing was performed using PCR-based protocols and Q20+ chemistry (Version 14 kits). Two technical replicates per mock sample were performed. DNA samples underwent human CpG-methylated DNA depletion using the NEBNext Microbiome DNA enrichment kit (New England Biolabs) prior library preparation using the Rapid PCR Barcoding kit 24 V 14 (SQK-RPB114.24) according to manufacturer’s instructions. RNA sequencing was performed using Rapid-Smart 9N(91). First, for annealing of the tagged random oligonucleotide, 10 µl of RNA was mixed with 1 μl of 2 μM RLB RT 9N oligo (TTTTTCGTGCGCCGCTTCAACNNNNNNNNN) and 1 μl 10 mM dNTPs. Mix was incubated for 5 min at 65°C, then cooled on ice. For cDNA synthesis and generation of double-tagged cDNA, 4 μlSuperScript IV First-strand Buffer, 1 μL 0.1 M DTT, 1 μl RNase OUT (Thermo Fisher Scientific), 1 μl 2 μM RLB TSO (GCTAATCATTGCTTTTTCGTGCGCCGCTTCAACATrGrGrG), and 1 μL SuperScript IV (Thermo Fisher Scientific) was mixed with the 12 μl annealed RNA. Reaction was incubated for 90 min at 42°C followed by 10 min at 70°C. 5 µl of double-tagged cDNA were used as input for the PCR step in the Rapid PCR Barcoding kit 24 V 14 (SQK-RPB114.24). From this step onwards, manufacturer’s instructions were followed.

All pre-PCR steps were carried out under an MSC class II cabinet and moved to a post-PCR area following amplification. Sequencing was performed using PromethION Flow cells (R.10.4.1) on a P2 solo device connected to a GridION. Real-time basecalling was performed in MinKnow Version 23.07.5 using the high-accuracy model. Samples were sequenced until a minimum output of 5 Gb per sample was obtained.

### Targeted Illumina sequencing with Twist Comprehensive Viral Research Panel

Targeted Illumina sequencing was performed on samples with a combined DNA + RNA background using the Twist Comprehensive Viral Research Panel (Twist Biosciences, 103550) following the Twist Biosciences Total Nucleic Acids Library Preparation EF Kit 2.0 for Viral Pathogen Detection and Characterization protocol. Two technical replicates per mock sample were processed other than for 60 and 600 gc/ml and the negative control, where four replicates were performed. Additional replicates were included to thoroughly test for potential cross-contamination and to assess potential sensitivity loss in low copy number samples when combined with high copy number samples in hybridisation-capture reactions.

First, cDNA synthesis was performed using ProtoScript II First strand synthesis kit (New England Biolabs, E6560) followed by the NEBNext Ultra Non-Directional Second Strand Synthesis module (New England Biolabs, E6111). 25ng of the double-stranded cDNA and dsDNA mix were used as input for adapter ligation, indexing and pre-capture amplification using the Twist Library preparation EF Kit 2.0 (Twist Biosciences, 104207 + 100573). All pre-PCR steps were carried out under an MSC class II cabinet until the indexing step was complete.

Following pre-capture amplification, indexed samples were pooled, for a total of 7 samples plus a negative control per hybridisation reaction. Hybridisation was performed overnight for 16 hours. Hybridisation targets were then captured with Streptavidin Binding Beads. At this step, samples were washed using the Twist Wash Buffers (Twist Biosciences, 104178) instead of the washing buffers V2 as per recommendation of the manufacturer. Post-capture amplification was performed on the enriched libraries (8 cycles). Final enriched libraries were quantified with Qubit high sensitivity kit and average peaks obtained with high sensitivity D1000 tapes. Samples were sequenced in equimolar pools using a NextSeq 2000 or a NovaSeq 6000 300 cycle kit (2 x 150 bp) depending on the number of samples processed. A minimum output of 5 Gb per sample was obtained (Supplementary Table 1).

### Databases for taxonomic classification

Since database composition has been shown to have a significant impact on the results of metagenomics (92), a common set of sequences was used to build the databases where possible. For the tools where it was possible to create a custom database (Kraken2 (58), Bracken (59), Dragen Metagenomics Pipeline (60), EPI2ME labs wf-metagenomics (61), metaMix (63), MEGAN-LR (62) and Kaiju (57)), a database was created based on the bacterial (complete genomes only), viral, fungal, protozoa and human nucleotide from Refseq (downloaded 6^th^ June 2023). Databases were built using the default parameters, other than for MEGAN-LR, where the recommended settings for ONT data described in (39) were used. A common set of taxonomy files downloaded from NCBI (31^st^ July 2023) were also used. Unplaced contigs were removed from the parasites and fungal nucleotide sequences prior to building the databases to reduce human contamination present in some of the reference sequences. It is not currently possible for the user to alter the databases for CZ ID (64) or One Codex (65), so the inbuilt databases were used.

### Read preprocessing and taxonomic classification

Reads were randomly subsampled from the raw output fastq files, using seqtk (93) sample for the Illumina data and a custom python script for the ONT data, to obtain 5 Gb for each sample across all the technologies.

Kraken2, Bracken and Kaiju were run through the nf-core Taxprofiler pipeline (94), which aims to provide a reproducible best-practice workflow for metagenomics analysis. As recommended, read preprocessing involving adaptor trimming and complexity filtering with fastp (95) was performed for Illumina but not ONT sequencing (96). Host removal was performed for both platforms by alignment to the human genome (version Ch38).

The reads obtained following preprocessing and host removal from the Taxprofiler pipeline were used as input to MEGAN-LR, run through the PB-metagenomics tools pipeline(97), with the adjustments for ONT sequencing recommended in (39).

For Illumina data processed with metaMix, a separate preprocessing pipeline was used for a more thorough removal of host reads. This involves read trimming using TrimGalore (98), followed by removal of human DNA/RNA and ribosomal RNA using alignment with both Bowtie2 (99) and BLAST (100,101). For the other classifiers, the time saved in classification was shorter than the time taken for the longer host removal pipeline, so a single alignment step is sufficient. For the ONT data, the output of the preprocessing and host removal from Taxprofiler was used as input. Reads were then aligned to the reference database with BLAST (nucleotide mode) and DIAMOND (102) (protein mode) before input to the metaMix R package. metaMix-fast is the first two steps of the metaMix R package, before the time-consuming MCMC step.

Raw reads were uploaded to CZ ID metagenomics workflow through the online interface. For CZ ID, the fields nr_count or nt_count were used for protein and nucleotide analyses respectively. Raw reads were also uploaded to the One Codex platform. The reads field from the reads field used for the analysis. For the Twist panel data, the Twist Comprehensive Viral Research Panel report was also run. Initially, this report failed to identify Reovirus, but this issue has since been rectified by One Codex.

Raw reads were also used as input into either Illumina’s Dragen Metagenomics Pipeline or ONT’s EPI2ME labs wf-metagenomics as appropriate. The same Kraken2 custom database as used for running Kraken2 through nf-core Taxprofiler was used, and both tools were run using the command-line interface.

### Alignment and sensitivity analysis

Reads were aligned to reference genomes downloaded from ACC using Bowtie2 (99) for the Illumina data and Minimap2 (103) for the ONT data, using the “very-sensitive” mode and the default parameters respectively. PCR duplicate reads were removed before calculating coverage and depth using samtools (104).

### Specificity analysis

To standardize the results between classifiers for comparison, taxonomic ranks were identified, organisms were classified as bacteria, viruses, fungi or other eukaryotes, and all reads assigned to taxonomic levels below species were assigned to the relevant species, using custom R scripts and the taxonomizr package (105). Where an organism was detected by both DNA and RNA sequencing, the result with the higher number of reads was retained. All analysis was performed in terms of reads rather than base pairs since not all classifiers output assignments by read, making it impossible to calculate base pair assignments for the ONT data. Read per million ratios and proportion of microbial reads were calculated and used to identify positive species as described in the supplementary information. False positive species were defined as species that were identified by the classifiers and were not present in the mock community or the positive controls. False positive viral species were classified according to host using the Virus-Host DB (106).

### Host transcriptomic analysis

Genes and transcripts were quantified using Kallisto(107) with human genome GRCh38.p14 downloaded from Gencode (108). Analysis was conducted in R using the tximport (109) and rtracklayer (110) packages. Spliced reads were identified by alignment to the human genome using STAR (111), using the presence of the CIGAR string to identify gapped alignments.

### Plots

Plots were produced in R using Tidyverse (112) packages or using Biorender.com.

## Data Availability

All data produced in the present study are available upon reasonable request to the authors.

## List of abbreviations

ONT: Oxford Nanopore Technologies
VRP: Viral Research Panel

## Declarations Ethics approval

This study used only commercially available human genetic material.

## Consent for publication

Not applicable

## Availability of data and code

The raw data produced in this study is available in the European Nucleotide Archives (ENA) with accession PRJEB74559. All scripts used for analysis are available at https://github.com/xxxxxx/viral-metagenomics-comparison. *(Reviewers please note that the data and code will be made public on publication.)*

## Author contributions

The study was designed by OETM, SM, JB, RW, SC, LMMB, NA, LF and SB. Laboratory work and data analysis were performed by SB, LF, NA, LMMB, TB, CV, CM, NS, LA, TB, SR, SG and OETM. SB, LF, JB, SM and OETM wrote and revised the manuscript. JRB, PS, HH, TG, CV and RW revised the manuscript. All authors read and approved the final manuscript.

## Acknowledgements

Not applicable

---

**Supplementary Figure 1:**
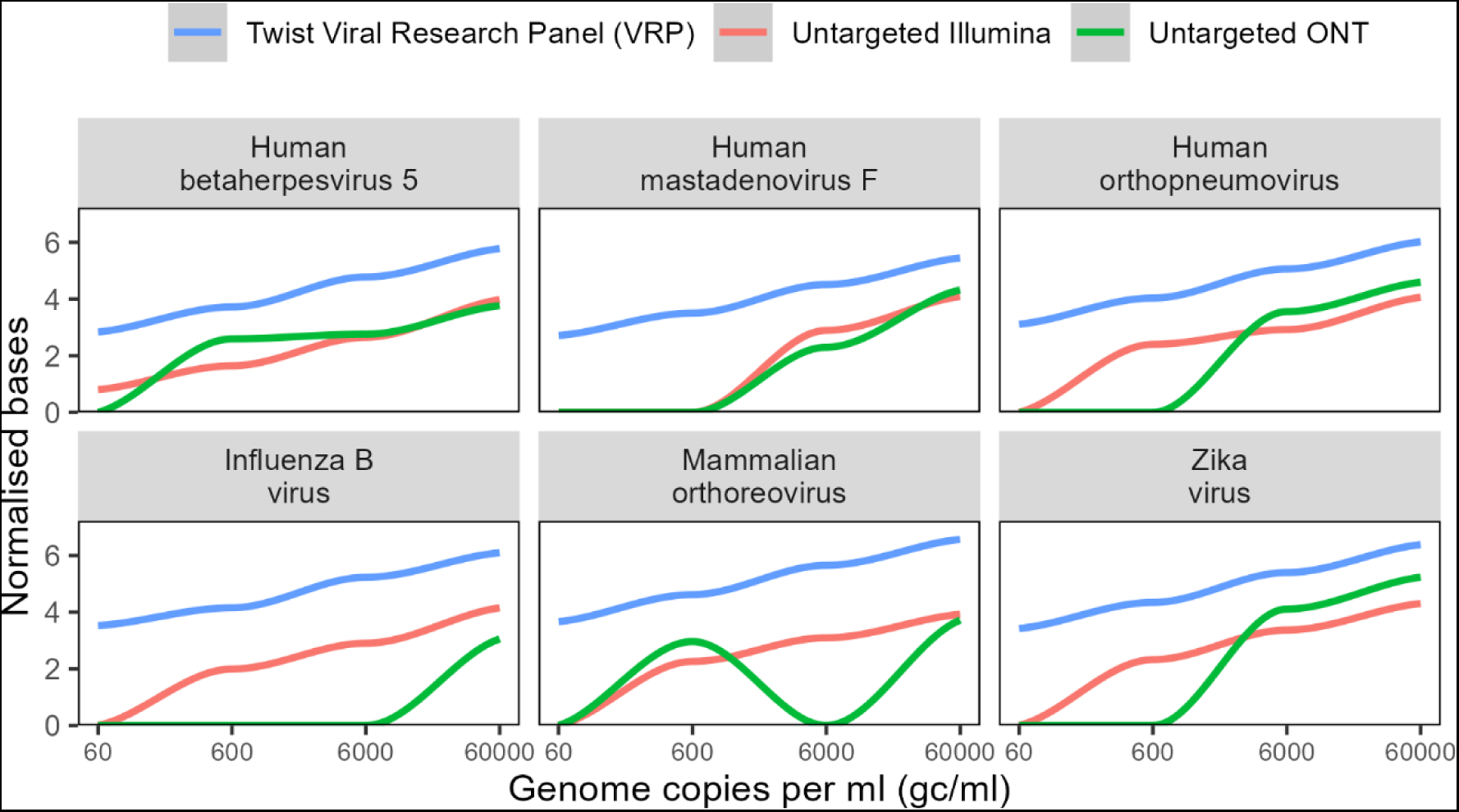
Sensitivity normalized by genome length. Normalised bases aligning to the genome of each species in the mock community, calculated as log10(base pairs * 10^4 / genome length).

**Supplementary Figure 2:**
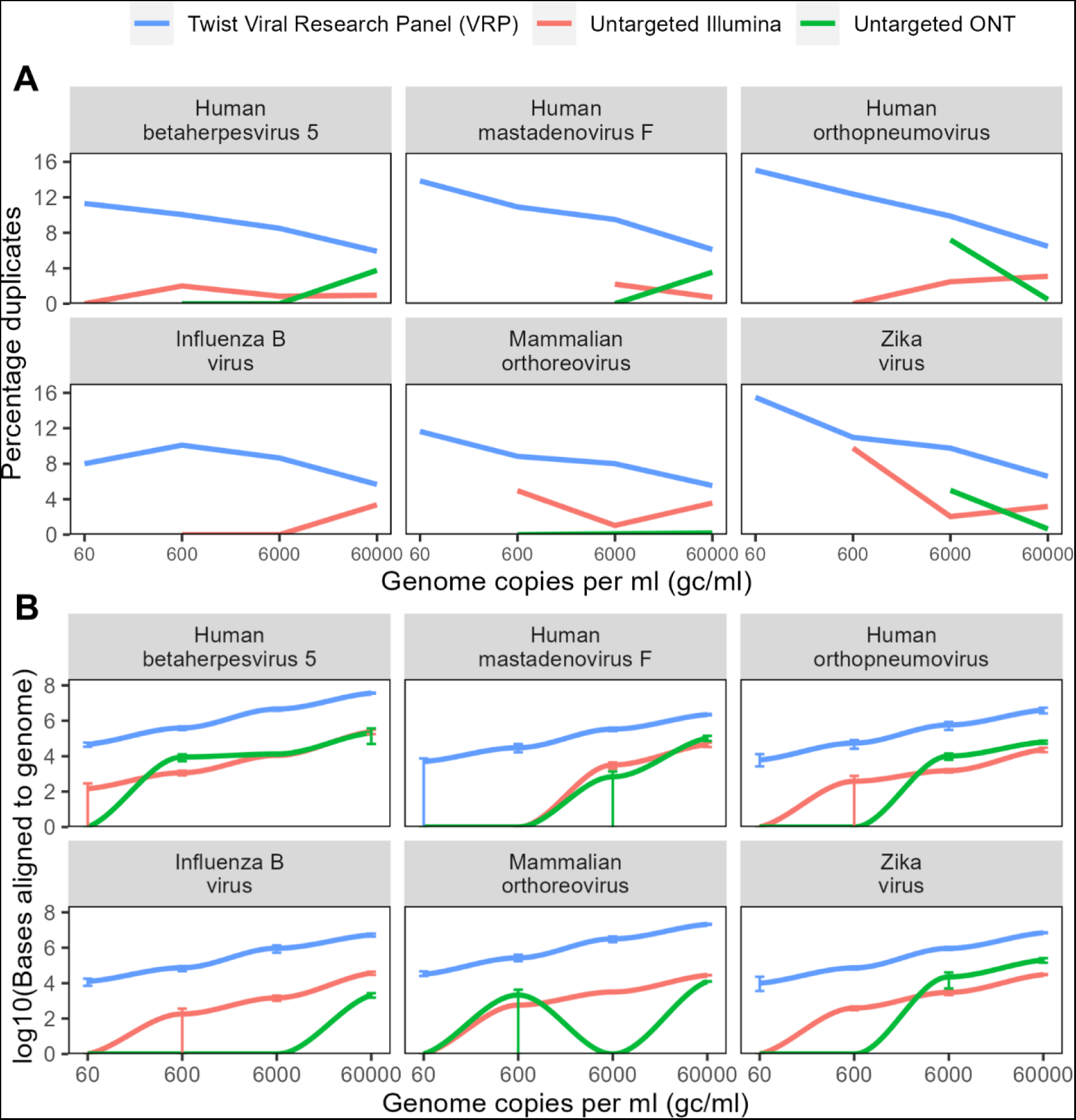
PCR duplicates. **A** Percentage of the total reads aligning to the genome of each species that were marked as duplicates by samtools markdup. **B** Figure 1B without PCR duplicated removed.

**Supplementary Figure 3:**
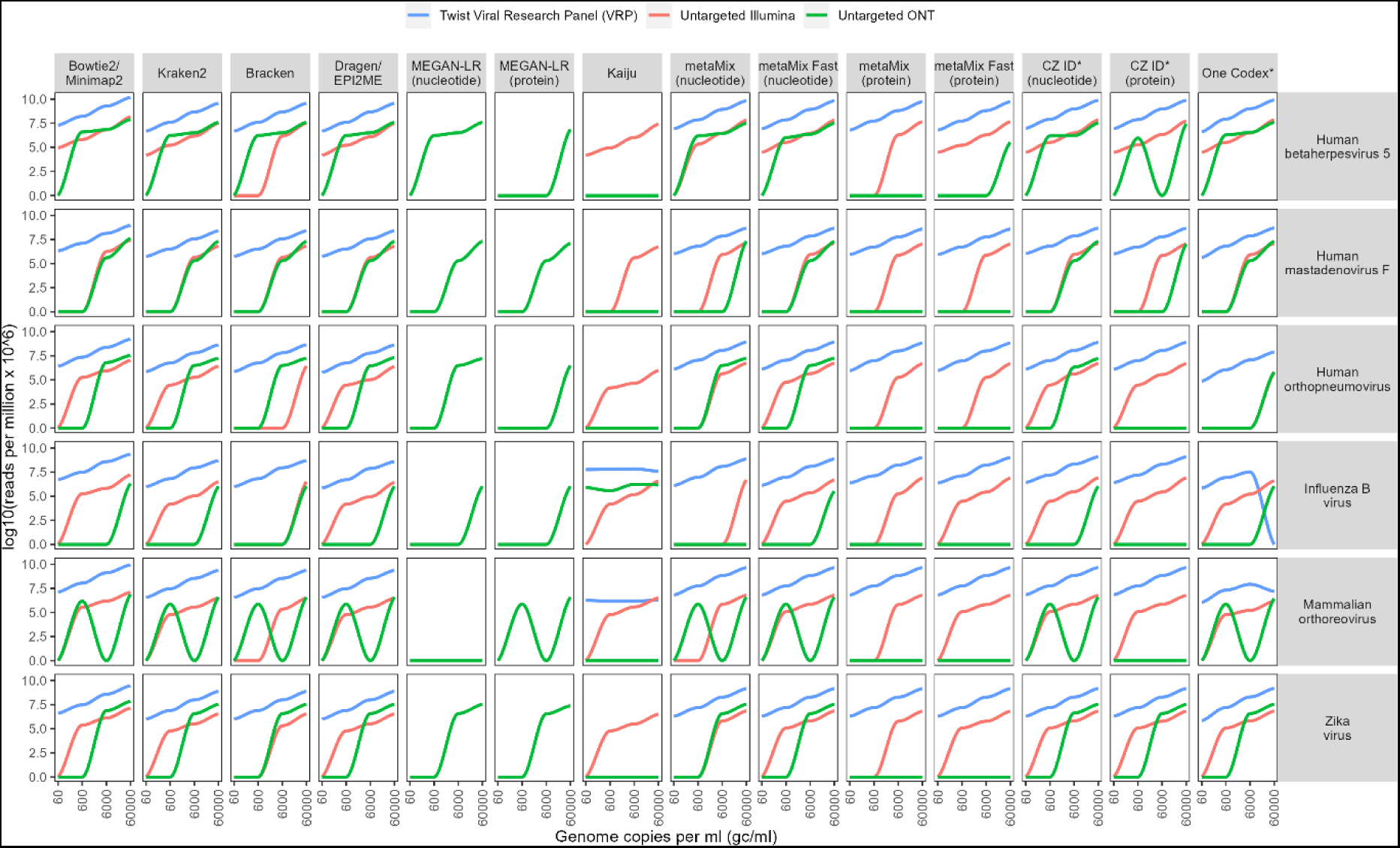
Reads classified. Number of reads assigned to each species in the mock community by classifiers. Each point shows the mean of at least two technical replicates.

**Supplementary Figure 4:**
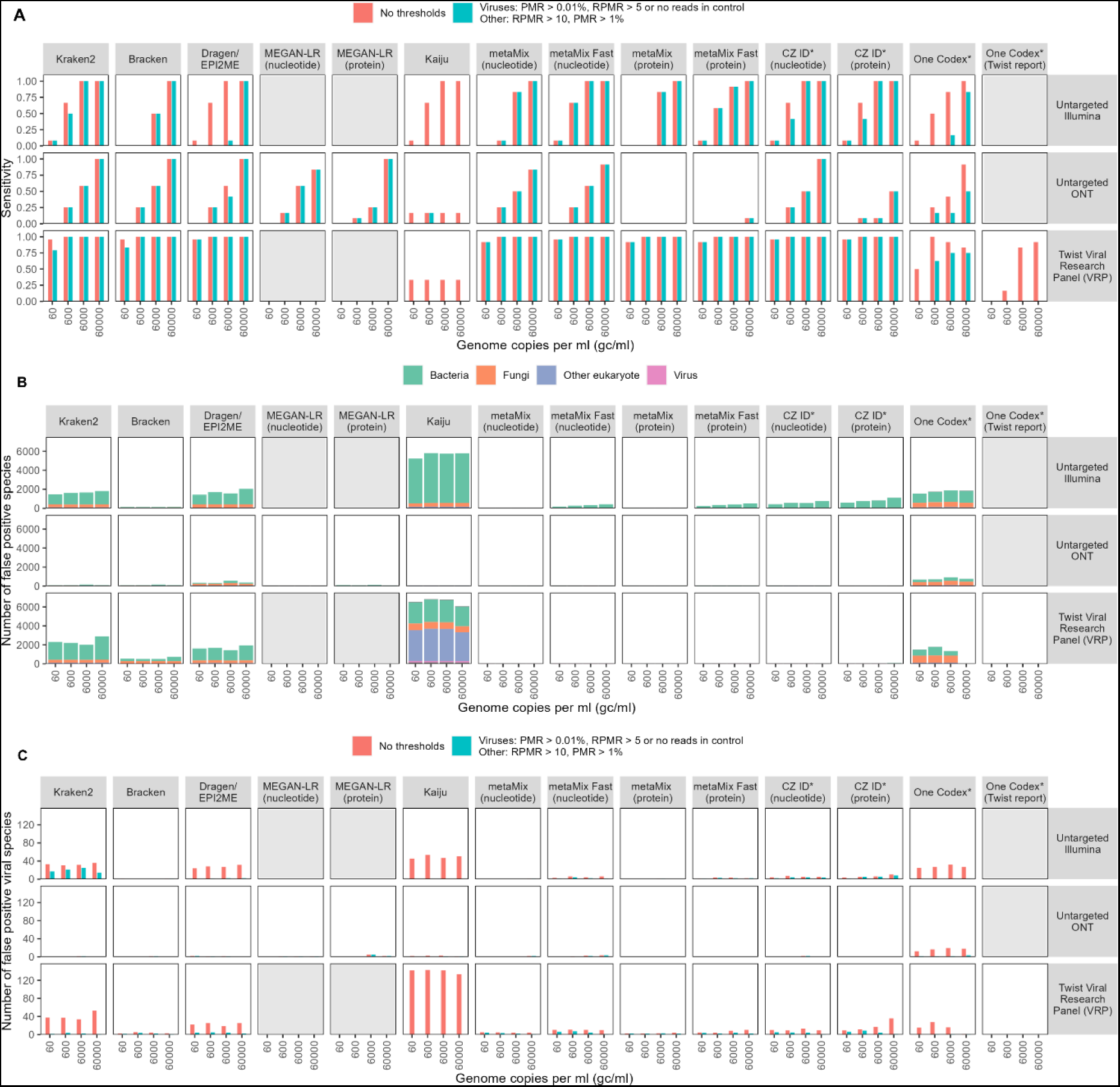
Sensitivity and specificity with nucleotide and protein-based classifiers. **A** Sensitivity to the species in the mock community before and after the application of thresholds, for eight different taxonomic classifiers, including their protein modes where relevant, by untargeted Illumina and ONT sequencing and capture probe enrichment with the Twist Biosciences Comprehensive Viral Research Panel followed by Illumina sequencing. MEGAN-LR and the One Codex Twist report are only designed for ONT and Twist sequencing respectively so were only run for these platforms. **B,C** Number of false positive species, defined as a species that is classified as positive but not present in the mock community **B** False positive species from the raw output of the taxonomic classifiers with no thresholds applied. **C** Comparison of the numbers of viral positive species identified before and after the application of thresholds. Genome copy numbers refer to an average across the viral species – see **Supplementary Table 3**. Each bar shows the mean of at least two technical replicates.

**Supplementary Figure 5:**
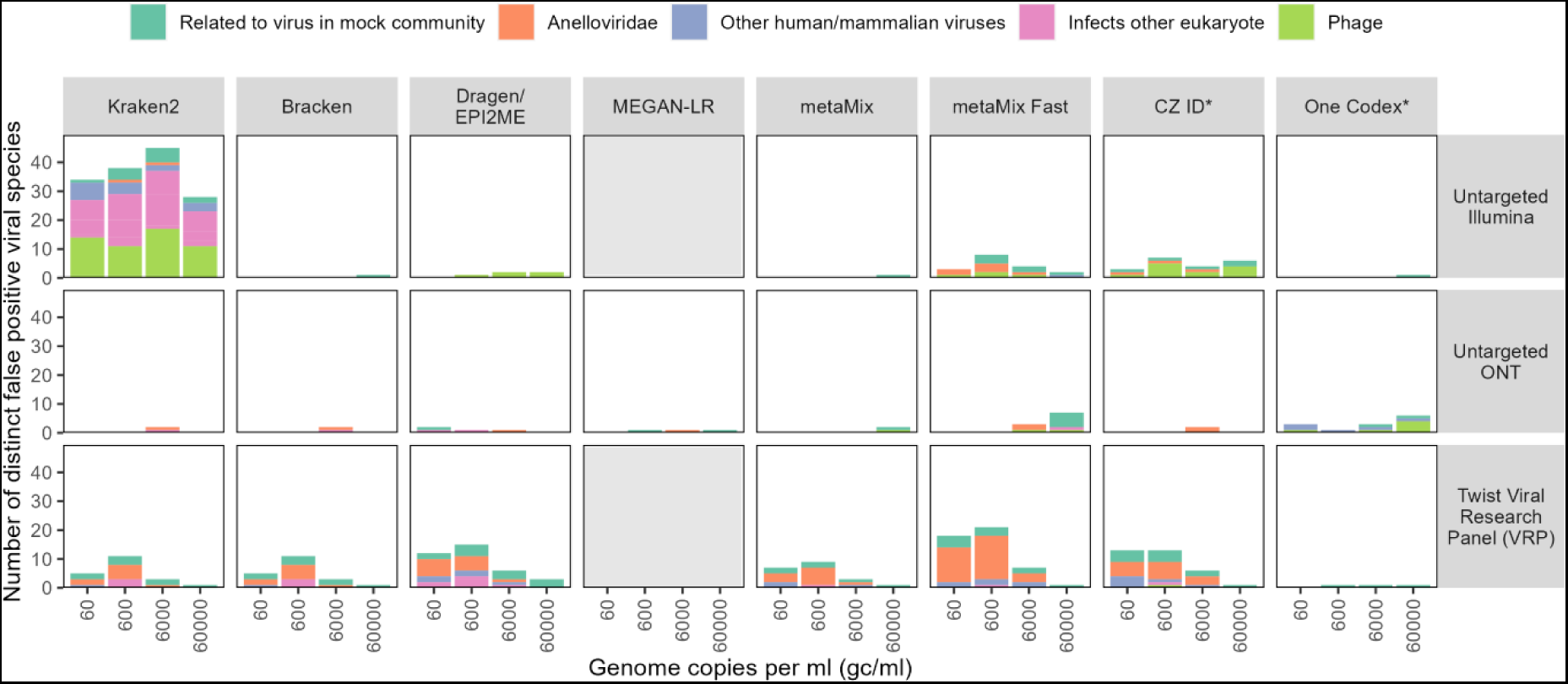
False positive viruses. Number of distinct false positive viral species, defined as a species that is classified as positive but not present in the mock community or positive controls, after application of thresholds described in Figure 3. Shows the number of distinct species across the 2-4 technical replicates, so totals may be slightly higher than in Figure 3c, which shows an average across technical replicates.

## References

1. Wu F, Zhao S, Yu B, Chen YM, Wang W, Song ZG, et al. A new coronavirus associated with human respiratory disease in China. Nature. 2020 Mar;579(7798):265–9.

2. Palacios G, Druce J, Du L, Tran T, Birch C, Briese T, et al. A New Arenavirus in a Cluster of Fatal Transplant-Associated Diseases. New England Journal of Medicine. 2008 Mar 6;358(10):991–8.

3. Quan PL, Wagner TA, Briese T, Torgerson TR, Hornig M, Tashmukhamedova A, et al. Astrovirus Encephalitis in Boy with X-linked Agammaglobulinemia. Emerg Infect Dis. 2010 Jun;16(6):918–25.

4. Wilson MR, Naccache SN, Samayoa E, Biagtan M, Bashir H, Yu G, et al. Actionable diagnosis of neuroleptospirosis by next-generation sequencing. N Engl J Med. 2014 Jun 19;370(25):2408–17.

5. Naccache SN, Peggs KS, Mattes FM, Phadke R, Garson JA, Grant P, et al. Diagnosis of Neuroinvasive Astrovirus Infection in an Immunocompromised Adult With Encephalitis by Unbiased Next-Generation Sequencing. Clinical Infectious Diseases. 2015 Mar 15;60(6):919–23.

6. Brown JR, Bharucha T, Breuer J. Encephalitis diagnosis using metagenomics: application of next generation sequencing for undiagnosed cases. Journal of Infection. 2018 Mar 1;76(3):225–40.

7. Brown JR, Morfopoulou S, Hubb J, Emmett WA, Ip W, Shah D, et al. Astrovirus VA1/HMO-C: An Increasingly Recognized Neurotropic Pathogen in Immunocompromised Patients. Clin Infect Dis. 2015 Mar 15;60(6):881–8.

8. Morfopoulou S, Mee ET, Connaughton SM, Brown JR, Gilmour K, Chong WK “Kling,” et al. Deep sequencing reveals persistence of cell-associated mumps vaccine virus in chronic encephalitis. Acta Neuropathol. 2017 Jan;133(1):139–47.

9. Morfopoulou S, Brown JR, Davies EG, Anderson G, Virasami A, Qasim W, et al. Human Coronavirus OC43 Associated with Fatal Encephalitis. N Engl J Med. 2016 Aug 4;375(5):497–8.

10. Wilson MR, Sample HA, Zorn KC, Arevalo S, Yu G, Neuhaus J, et al. Clinical Metagenomic Sequencing for Diagnosis of Meningitis and Encephalitis. N Engl J Med. 2019 Jun 13;380(24):2327–40.

11. Penner J, Hassell J, Brown JR, Mankad K, Storey N, Atkinson L, et al. Translating metagenomics into clinical practice for complex paediatric neurological presentations. Journal of Infection. 2023 Nov 1;87(5):451–8.

12. Charalampous T, Alcolea-Medina A, Snell LB, Alder C, Tan M, Williams TGS, et al. Routine Metagenomics Service for ICU Patients with Respiratory Infection. Am J Respir Crit Care Med. 2024 Jan 15;209(2):164–74.

13. Gu W, Deng X, Lee M, Sucu YD, Arevalo S, Stryke D, et al. Rapid pathogen detection by metagenomic next-generation sequencing of infected body fluids. Nat Med. 2021 Jan;27(1):115–24.

14. Greninger AL, Naccache SN, Federman S, Yu G, Mbala P, Bres V, et al. Rapid metagenomic identification of viral pathogens in clinical samples by real-time nanopore sequencing analysis. Genome Medicine. 2015 Sep 29;7(1):99.

15. Pendleton KM, Erb-Downward JR, Bao Y, Branton WR, Falkowski NR, Newton DW, et al. Rapid Pathogen Identification in Bacterial Pneumonia Using Real-Time Metagenomics. Am J Respir Crit Care Med. 2017 Dec 15;196(12):1610–2.

16. Charalampous T, Alcolea-Medina A, Snell LB, Williams TGS, Batra R, Alder C, et al. Evaluating the potential for respiratory metagenomics to improve treatment of secondary infection and detection of nosocomial transmission on expanded COVID-19 intensive care units. Genome Medicine. 2021 Nov 17;13(1):182.

17. Charalampous T, Kay GL, Richardson H, Aydin A, Baldan R, Jeanes C, et al. Nanopore metagenomics enables rapid clinical diagnosis of bacterial lower respiratory infection. Nat Biotechnol. 2019 Jul;37(7):783–92.

18. Lin Q, Yao Y, Li X, Zhang S, Guo H, Ma X, et al. The application of nanopore targeted sequencing for pathogen diagnosis in bronchoalveolar lavage fluid of patients with pneumonia: a prospective multicenter study. Infectious Diseases. 2024 Feb 1;56(2):128–37.

19. Wylie TN, Wylie KM, Herter BN, Storch GA. Enhanced virome sequencing using targeted sequence capture. Genome Res. 2015 Dec;25(12):1910–20.

20. Briese T, Kapoor A, Mishra N, Jain K, Kumar A, Jabado OJ, et al. Virome Capture Sequencing Enables Sensitive Viral Diagnosis and Comprehensive Virome Analysis. mBio. 2015 Sep 22;6(5):10.1128/mbio.01491-15.

21. Kapel N, Kalimeris E, Lumley S, Decano A, Rodger G, Lopes Alves M, et al. Evaluation of sequence hybridization for respiratory viruses using the Twist Bioscience Respiratory Virus Research panel and the OneCodex Respiratory Virus sequence analysis workflow. Microbial Genomics. 2023;9(9):001103.

22. Deng X, Achari A, Federman S, Yu G, Somasekar S, Bártolo I, et al. Metagenomic sequencing with spiked primer enrichment for viral diagnostics and genomic surveillance. Nat Microbiol. 2020 Mar;5(3):443–54.

23. Jansen SA, Nijhuis W, Leavis HL, Riezebos-Brilman A, Lindemans CA, Schuurman R. Targeted Sequence Capture Metagenomics (ViroCap) to Detect Viruses in Stool Samples of Hematopoietic Stem Cell Transplantation Patients. Biology of Blood and Marrow Transplantation. 2020 Mar 1;26(3, Supplement):S174–5.

24. Wylie KM, Wylie TN, Buller R, Herter B, Cannella MT, Storch GA. Detection of Viruses in Clinical Samples by Use of Metagenomic Sequencing and Targeted Sequence Capture. Journal of Clinical Microbiology. 2018 Nov 27;56(12):10.1128/jcm.01123-18.

25. Sczyrba A, Hofmann P, Belmann P, Koslicki D, Janssen S, Dröge J, et al. Critical Assessment of Metagenome Interpretation—a benchmark of metagenomics software. Nat Methods. 2017 Nov;14(11):1063–71.

26. Meyer F, Fritz A, Deng ZL, Koslicki D, Lesker TR, Gurevich A, et al. Critical Assessment of Metagenome Interpretation: the second round of challenges. Nat Methods. 2022 Apr;19(4):429–40.

27. Horiba K, Torii Y, Aizawa Y, Yamaguchi M, Haruta K, Okumura T, et al. Performance of Nanopore and Illumina Metagenomic Sequencing for Pathogen Detection and Transcriptome Analysis in Infantile Central Nervous System Infections. Open Forum Infectious Diseases. 2022 Oct 1;9(10):ofac504.

28. Gehrig JL, Portik DM, Driscoll MD, Jackson E, Chakraborty S, Gratalo D, et al. Finding the right fit: evaluation of short-read and long-read sequencing approaches to maximize the utility of clinical microbiome data. Microb Genom. 2022 Mar 18;8(3):000794.

29. Sevim V, Lee J, Egan R, Clum A, Hundley H, Lee J, et al. Shotgun metagenome data of a defined mock community using Oxford Nanopore, PacBio and Illumina technologies. Sci Data. 2019 Nov 26;6(1):285.

30. Meslier V, Quinquis B, Da Silva K, Plaza Oñate F, Pons N, Roume H, et al. Benchmarking second and third-generation sequencing platforms for microbial metagenomics. Sci Data. 2022 Nov 11;9(1):694.

31. Mori H, Kato T, Ozawa H, Sakamoto M, Murakami T, Taylor TD, et al. Assessment of metagenomic workflows using a newly constructed human gut microbiome mock community. DNA Research. 2023 Jun 1;30(3):dsad010.

32. Nakamura A, Komatsu M. Performance evaluation of whole genome metagenomics sequencing with the MinION nanopore sequencer: Microbial community analysis and antimicrobial resistance gene detection. J Microbiol Methods. 2023 Mar;206:106688.

33. Pearman WS, Freed NE, Silander OK. Testing the advantages and disadvantages of short- and long-read eukaryotic metagenomics using simulated reads. BMC Bioinformatics. 2020 May 29;21(1):220.

34. Cook R, Brown N, Rihtman B, Michniewski S, Redgwell T, Clokie M, et al. The long and short of it: benchmarking viromics using Illumina, Nanopore and PacBio sequencing technologies. Microb Genom. 2024 Feb 20;10(2):001198.

35. Cadenas-Castrejón E, Verleyen J, Boukadida C, Díaz-González L, Taboada B. Evaluation of tools for taxonomic classification of viruses. Briefings in Functional Genomics. 2023 Jan 1;22(1):31–41.

36. de Vries JJC, Brown JR, Fischer N, Sidorov IA, Morfopoulou S, Huang J, et al. Benchmark of thirteen bioinformatic pipelines for metagenomic virus diagnostics using datasets from clinical samples. Journal of Clinical Virology. 2021 Aug 1;141:104908.

37. Ye SH, Siddle KJ, Park DJ, Sabeti PC. Benchmarking Metagenomics Tools for Taxonomic Classification. Cell. 2019 Aug 8;178(4):779–94.

38. Govender KN, Eyre DW. Benchmarking taxonomic classifiers with Illumina and Nanopore sequence data for clinical metagenomic diagnostic applications. Microb Genom. 2022 Oct;8(10).

39. Portik DM, Brown CT, Pierce-Ward NT. Evaluation of taxonomic classification and profiling methods for long-read shotgun metagenomic sequencing datasets. BMC Bioinformatics. 2022 Dec 13;23(1):541.

40. Marić J, Križanović K, Riondet S, Nagarajan N, Šikić M. Comparative analysis of metagenomic classifiers for long-read sequencing datasets. BMC Bioinformatics. 2024 Jan 11;25(1):15.

41. Dunn G, Klapsa D, Wilton T, Stone L, Minor PD, Martin J. Twenty-Eight Years of Poliovirus Replication in an Immunodeficient Individual: Impact on the Global Polio Eradication Initiative. PLoS Pathog. 2015 Aug 27;11(8):e1005114.

42. Kaiser L, Aubert JD, Pache JC, Deffernez C, Rochat T, Garbino J, et al. Chronic Rhinoviral Infection in Lung Transplant Recipients. Am J Respir Crit Care Med. 2006 Dec 15;174(12):1392–9.

43. Pinsky BA, Mix S, Rowe J, Ikemoto S, Baron EJ. Long-term Shedding of Influenza A Virus in Stool of Immunocompromised Child. Emerg Infect Dis. 2010 Jul;16(7):1165–7.

44. Razonable RR, Inoue N, Pinninti SG, Boppana SB, Lazzarotto T, Gabrielli L, et al. Clinical Diagnostic Testing for Human Cytomegalovirus Infections. J Infect Dis. 2020 Mar 15;221(Suppl 1):S74–85.

45. Boggild AK, Geduld J, Libman M, Yansouni CP, McCarthy AE, Hajek J, et al. Surveillance report of Zika virus among Canadian travellers returning from the Americas. CMAJ. 2017 Mar 6;189(9):E334–40.

46. Jerome H, Taylor C, Sreenu VB, Klymenko T, Filipe ADS, Jackson C, et al. Metagenomic next-generation sequencing aids the diagnosis of viral infections in febrile returning travellers. Journal of Infection. 2019 Oct 1;79(4):383–8.

47. Lopez-Labrador FX, Huber M, Sidorov IA, Brown JR, Cuypers L, Laenen L, et al. Multicenter benchmarking of short and long read wet lab protocols for clinical viral metagenomics [Internet]. medRxiv; 2024 [cited 2024 Mar 18]. p. 2024.01.14.24301284. Available from: https://www.medrxiv.org/content/10.1101/2024.01.14.24301284v1

48. Unified metagenomic method for rapid detection of bacteria, fungi and viruses in clinical samples [Internet]. 2023 [cited 2023 Dec 22]. Available from: https://www.researchsquare.com

49. Yap M, Feehily C, Walsh CJ, Fenelon M, Murphy EF, McAuliffe FM, et al. Evaluation of methods for the reduction of contaminating host reads when performing shotgun metagenomic sequencing of the milk microbiome. Sci Rep. 2020 Dec 10;10(1):21665.

50. Kalantar KL, Neyton L, Abdelghany M, Mick E, Jauregui A, Caldera S, et al. Integrated host-microbe plasma metagenomics for sepsis diagnosis in a prospective cohort of critically ill adults. Nat Microbiol. 2022 Nov;7(11):1805–16.

51. Langelier C, Kalantar KL, Moazed F, Wilson MR, Crawford ED, Deiss T, et al. Integrating host response and unbiased microbe detection for lower respiratory tract infection diagnosis in critically ill adults. Proceedings of the National Academy of Sciences. 2018 Dec 26;115(52):E12353–62.

52. Mick E, Tsitsiklis A, Kamm J, Kalantar KL, Caldera S, Lyden A, et al. Integrated host/microbe metagenomics enables accurate lower respiratory tract infection diagnosis in critically ill children. J Clin Invest. 133(7):e165904.

53. Ramachandran PS, Ramesh A, Creswell FV, Wapniarski A, Narendra R, Quinn CM, et al. Integrating central nervous system metagenomics and host response for diagnosis of tuberculosis meningitis and its mimics. Nat Commun. 2022 Mar 30;13(1):1675.

54. Chiu CY, Miller SA. Clinical metagenomics. Nat Rev Genet. 2019 Jun;20(6):341–55.

55. Schuele L, Cassidy H, Peker N, Rossen JWA, Couto N. Future potential of metagenomics in microbiology laboratories. Expert Review of Molecular Diagnostics. 2021 Dec 2;21(12):1273–85.

56. One Codex Database | One Codex Docs [Internet]. [cited 2024 Mar 19]. Available from: http://docs.onecodex.com/en/articles/3761205-one-codex-database

57. Menzel P, Ng KL, Krogh A. Fast and sensitive taxonomic classification for metagenomics with Kaiju. Nat Commun. 2016 Apr 13;7(1):11257.

58. Wood DE, Lu J, Langmead B. Improved metagenomic analysis with Kraken 2. Genome Biology. 2019 Nov 28;20(1):257.

59. Lu J, Breitwieser FP, Thielen P, Salzberg SL. Bracken: estimating species abundance in metagenomics data. PeerJ Comput Sci. 2017 Jan 2;3:e104.

60. DRAGEN Metagenomics Pipeline [Internet]. [cited 2023 Oct 4]. Available from: https://emea.illumina.com/products/by-type/informatics-products/basespace-sequence-hub/apps/dragen-metagenomics-pipeline.html

61. wf-metagenomics [Internet]. EPI2ME Labs; 2023 [cited 2023 Aug 21]. Available from: https://github.com/epi2me-labs/wf-metagenomics

62. Huson DH, Albrecht B, Bağcı C, Bessarab I, Górska A, Jolic D, et al. MEGAN-LR: new algorithms allow accurate binning and easy interactive exploration of metagenomic long reads and contigs. Biology Direct. 2018 Apr 20;13(1):6.

63. Morfopoulou S, Plagnol V. Bayesian mixture analysis for metagenomic community profiling. Bioinformatics. 2015 Sep 15;31(18):2930–8.

64. Kalantar KL, Carvalho T, de Bourcy CFA, Dimitrov B, Dingle G, Egger R, et al. IDseq—An open source cloud-based pipeline and analysis service for metagenomic pathogen detection and monitoring. GigaScience. 2020 Oct 20;9(10):giaa111.

65. Minot SS, Krumm N, Greenfield NB. One Codex: A Sensitive and Accurate Data Platform for Genomic Microbial Identification [Internet]. bioRxiv; 2015 [cited 2024 Mar 4]. p. 027607. Available from: https://www.biorxiv.org/content/10.1101/027607v2

66. Average Sterling exchange rate: US Dollar XUMAUSS - Office for National Statistics [Internet]. [cited 2024 Mar 18]. Available from: https://www.ons.gov.uk/economy/nationalaccounts/balanceofpayments/timeseries/auss/mret

67. Schulz E, Grumaz S, Hatzl S, Gornicec M, Valentin T, Huber-Kraßnitzer B, et al. Pathogen Detection by Metagenomic Next-Generation Sequencing During Neutropenic Fever in Patients With Hematological Malignancies. Open Forum Infectious Diseases. 2022 Aug 1;9(8):ofac393.

68. Hogan CA, Yang S, Garner OB, Green DA, Gomez CA, Dien Bard J, et al. Clinical Impact of Metagenomic Next-Generation Sequencing of Plasma Cell-Free DNA for the Diagnosis of Infectious Diseases: A Multicenter Retrospective Cohort Study. Clinical Infectious Diseases. 2021 Jan 15;72(2):239–45.

69. Wilke J, Ramchandar N, Cannavino C, Pong A, Tremoulet A, Padua LT, et al. Clinical application of cell-free next-generation sequencing for infectious diseases at a tertiary children’s hospital. BMC Infectious Diseases. 2021 Jun 11;21(1):552.

70. Klenner J, Kohl C, Dabrowski PW, Nitsche A. Comparing Viral Metagenomic Extraction Methods. Current Issues in Molecular Biology. 2017 Oct;24(1):59–70.

71. Zhao X, Ge Y, Zhang Y, Zhang W, Hu H, Li L, et al. Pathogen Diagnosis Value of Nanopore Sequencing in Severe Hospital-Acquired Pneumonia Patients. IDR. 2023 May 26;16:3293–303.

72. Low L, Nakamichi K, Akileswaran L, Lee CS, Lee AY, Moussa G, et al. Deep Metagenomic Sequencing for Endophthalmitis Pathogen Detection Using a Nanopore Platform. American Journal of Ophthalmology. 2022 Oct 1;242:243–51.

73. Yamaguchi M, Horiba K, Haruta K, Takeuchi S, Suzuki T, Torii Y, et al. Utility of nanopore sequencing for detecting pathogens in bronchoalveolar lavage fluid from pediatric patients with respiratory failure. Journal of Clinical Virology Plus. 2023 Jun 1;3(2):100154.

74. Zhao N, Cao J, Xu J, Liu B, Liu B, Chen D, et al. Targeting RNA with Next- and Third-Generation Sequencing Improves Pathogen Identification in Clinical Samples. Adv Sci (Weinh). 2021 Oct 23;8(23):2102593.

75. Jia X, Hu L, Wu M, Ling Y, Wang W, Lu H, et al. A streamlined clinical metagenomic sequencing protocol for rapid pathogen identification. Sci Rep. 2021 Feb 23;11(1):4405.

76. Pichler I, Schmutz S, Ziltener G, Zaheri M, Kufner V, Trkola A, et al. Rapid and sensitive single-sample viral metagenomics using Nanopore Flongle sequencing. Journal of Virological Methods. 2023 Oct 1;320:114784.

77. Zhang J, Gao L, Zhu C, Jin J, Song C, Dong H, et al. Clinical value of metagenomic next-generation sequencing by Illumina and Nanopore for the detection of pathogens in bronchoalveolar lavage fluid in suspected community-acquired pneumonia patients. Front Cell Infect Microbiol. 2022 Sep 27;12:1021320.

78. Nicholls SM, Quick JC, Tang S, Loman NJ. Ultra-deep, long-read nanopore sequencing of mock microbial community standards. GigaScience. 2019 May 1;8(5):giz043.

79. Culviner PH, Guegler CK, Laub MT. A Simple, Cost-Effective, and Robust Method for rRNA Depletion in RNA-Sequencing Studies. mBio. 2020 Apr 21;11(2):10.1128/mbio.00010-20.

80. Street TL, Barker L, Sanderson ND, Kavanagh J, Hoosdally S, Cole K, et al. Optimizing DNA Extraction Methods for Nanopore Sequencing of Neisseria gonorrhoeae Directly from Urine Samples. J Clin Microbiol. 2020 Feb 24;58(3):e01822–19.

81. Edgeworth JD. Respiratory metagenomics: route to routine service. Current Opinion in Infectious Diseases. 2023 Apr;36(2):115.

82. Comprehensive Viral Research Panel - Twist Bioscience [Internet]. [cited 2024 Feb 16]. Available from: https://www.twistbioscience.com/products/ngs/fixed-panels/comprehensive-viral-research-panel

83. Ogunbayo AE, Sabiu S, Nyaga MM. Evaluation of extraction and enrichment methods for recovery of respiratory RNA viruses in a metagenomics approach. Journal of Virological Methods. 2023 Apr 1;314:114677.

84. Gand M, Bloemen B, Vanneste K, Roosens NHC, De Keersmaecker SCJ. Comparison of 6 DNA extraction methods for isolation of high yield of high molecular weight DNA suitable for shotgun metagenomics Nanopore sequencing to detect bacteria. BMC Genomics. 2023 Aug 4;24(1):438.

85. Rios M, Daniel S, Chancey C, Hewlett IK, Stramer SL. West Nile Virus Adheres to Human Red Blood Cells in Whole Blood. Clinical Infectious Diseases. 2007 Jul 15;45(2):181–6.

86. Voermans JJC, Suzan D. Pas, Linden A van der, GeurtsvanKessel C, Koopmans M, Eijk A van der, et al. Whole-Blood Testing for Diagnosis of Acute Zika Virus Infections in Routine Diagnostic Setting - Volume 25, Number 7—July 2019 - Emerging Infectious Diseases journal - CDC. [cited 2024 Mar 12]; Available from: https://wwwnc.cdc.gov/eid/article/25/7/18-2000_article

87. Dollard SC, Roback JD, Gunthel C, Amin MM, Barclay S, Patrick E, et al. Measurements of human herpesvirus 8 viral load in blood before and after leukoreduction filtration. Transfusion. 2013 Oct;53(10):2164–7.

88. Hasan MR, Rawat A, Tang P, Jithesh PV, Thomas E, Tan R, et al. Depletion of Human DNA in Spiked Clinical Specimens for Improvement of Sensitivity of Pathogen Detection by Next-Generation Sequencing. Journal of Clinical Microbiology. 2016 Mar 25;54(4):919–27.

89. Virome Virus Mix - MSA-2008 | ATCC [Internet]. [cited 2023 Jan 27]. Available from: https://www.atcc.org/products/msa-2008

90. Atkinson L, Lee JCD, Lennon A, Shah D, Storey N, Morfopoulou S, et al. Untargeted metagenomics protocol for the diagnosis of infection from CSF and tissue from sterile sites. Heliyon. 2023 Sep 1;9(9):e19854.

91. Claro IM, Ramundo MS, Coletti TM, da Silva CAM, Valenca IN, Candido DS, et al. Rapid viral metagenomics using SMART-9N amplification and nanopore sequencing. Wellcome Open Res. 2021;6:241.

92. Breitwieser FP, Lu J, Salzberg SL. A review of methods and databases for metagenomic classification and assembly. Briefings in Bioinformatics. 2019 Jul 19;20(4):1125–36.

93. Li H. lh3/seqtk [Internet]. 2021 [cited 2021 Feb 25]. Available from: https://github.com/lh3/seqtk

94. Ewels PA, Peltzer A, Fillinger S, Patel H, Alneberg J, Wilm A, et al. The nf-core framework for community-curated bioinformatics pipelines. Nat Biotechnol. 2020 Mar;38(3):276–8.

95. Chen S, Zhou Y, Chen Y, Gu J. fastp: an ultra-fast all-in-one FASTQ preprocessor. Bioinformatics. 2018 Sep 1;34(17):i884–90.

96. taxprofiler: Introduction [Internet]. [cited 2023 Aug 11]. Available from: https://nf-co.re/taxprofiler/1.0.1.html

97. PB-metagenomics-tools [Internet]. PacBio; 2023 [cited 2023 Aug 11]. Available from: https://github.com/PacificBiosciences/pb-metagenomics-tools

98. Krueger F, James F, Ewels P, Afyounian E, Schuster-Boeckler B. FelixKrueger/TrimGalore: v0.6.7 - DOI via Zenodo [Internet]. Zenodo; 2021 [cited 2022 Jun 14]. Available from: https://zenodo.org/record/5127899

99. Langmead B, Salzberg SL. Fast gapped-read alignment with Bowtie 2. Nat Methods. 2012 Mar 4;9(4):357–9.

100. Camacho C, Coulouris G, Avagyan V, Ma N, Papadopoulos J, Bealer K, et al. BLAST+: architecture and applications. BMC Bioinformatics. 2009 Dec 15;10(1):421.

101. Morfopoulou S. smorfopoulou/clinical_metagenomics [Internet]. 2023 [cited 2024 Mar 26]. Available from: https://github.com/smorfopoulou/clinical_metagenomics

102. Buchfink B, Xie C, Huson DH. Fast and sensitive protein alignment using DIAMOND. Nat Methods. 2015 Jan;12(1):59–60.

103. Li H. Minimap2: pairwise alignment for nucleotide sequences. Bioinformatics. 2018 Sep 15;34(18):3094–100.

104. Li H, Handsaker B, Wysoker A, Fennell T, Ruan J, Homer N, et al. The Sequence Alignment/Map format and SAMtools. Bioinformatics. 2009 Aug 15;25(16):2078–9.

105. Sherrill-Mix S. Taxonomizr [Internet]. 2023 [cited 2023 Jun 2]. Available from: https://github.com/sherrillmix/taxonomizr

106. Mihara T, Nishimura Y, Shimizu Y, Nishiyama H, Yoshikawa G, Uehara H, et al. Linking Virus Genomes with Host Taxonomy. Viruses. 2016 Mar;8(3):66.

107. Bray NL, Pimentel H, Melsted P, Pachter L. Near-optimal probabilistic RNA-seq quantification. Nat Biotechnol. 2016 May;34(5):525–7.

108. Frankish A, Diekhans M, Jungreis I, Lagarde J, Loveland JE, Mudge JM, et al. GENCODE 2021. Nucleic Acids Res. 2020 Dec 3;49(D1):D916–23.

109. Soneson C, Love MI, Robinson MD. Differential analyses for RNA-seq: transcript-level estimates improve gene-level inferences. F1000Res. 2016 Feb 29;4:1521.

110. Lawrence M, Gentleman R, Carey V. rtracklayer: an R package for interfacing with genome browsers. Bioinformatics. 2009 Jul 15;25(14):1841–2.

111. Dobin A, Davis CA, Schlesinger F, Drenkow J, Zaleski C, Jha S, et al. STAR: ultrafast universal RNA-seq aligner. Bioinformatics. 2013 Jan;29(1):15–21.

112. Wickham H, Averick M, Bryan J, Chang W, McGowan L, François R, et al. Welcome to the Tidyverse. JOSS. 2019 Nov 21;4(43):1686.

