## Supplementary Information for "Evaluating metagenomics and targeted approaches for diagnosis and surveillance of viruses"

### Clinical metagenomics for detection of viruses using short-read, long-read and targeted approaches

[1]Infection, Immunity and Inflammation Department, Great Ormond Street Institute of Child Health, University College London, London, UK, [2]Genetics and Genomic Medicine Department, Great Ormond Street Institute of Child Health, University College London, London, UK, [3]Department of Microbiology, Virology and Infection Prevention & Control, Great Ormond Street Hospital for Children NHS Foundation Trust, London, UK, [4]Nuffield Department of Medicine, University of Oxford, Oxford, UK, [5]Radcliffe Department of Medicine, University of Oxford, Oxford, UK, [6]Division of Infection and Immunity, University College London, London, UK, [7]Microbiology Services, NHS Blood and Transplant, Colindale, UK, [8]Sydney Infectious Diseases Institute, Faculty of Medicine and Health, University of Sydney, Sydney, Australia, [9]Section for Paediatrics, Department of Infectious Diseases, Faculty of Medicine, London, UK

* These authors contributed equally

[c] Corresponding author. These authors contributed equally

#### Determination of thresholds for calling positive viral species

Identification of false positive species is common in metagenomics analysis, with contamination arising at various stages of the laboratory and bioinformatics protocols (1). Therefore, approaches to distinguish true and false positives post-analysis are required. Most metagenomics protocols for clinical diagnostics implement thresholds based on read counts (2,3) or genome coverage (4).

We initially tested the classifiers with no thresholds applied. Some classifiers detected a high number of false positive species, mainly bacteria and fungi (**SI Figure 1A**). There were few false positive viral species detected by ONT sequencing by any classifier (**SI Figure 1B**), meaning that it may be possible to only perform a comparison with the negative control for these samples. metaMix and Bracken generally identified a low number of false positive viral species for the Illumina data, with other classifiers finding more (**SI Figure 1B**). For some classifiers, use of thresholds might reduce the number of false positive species whilst retaining sensitivity.

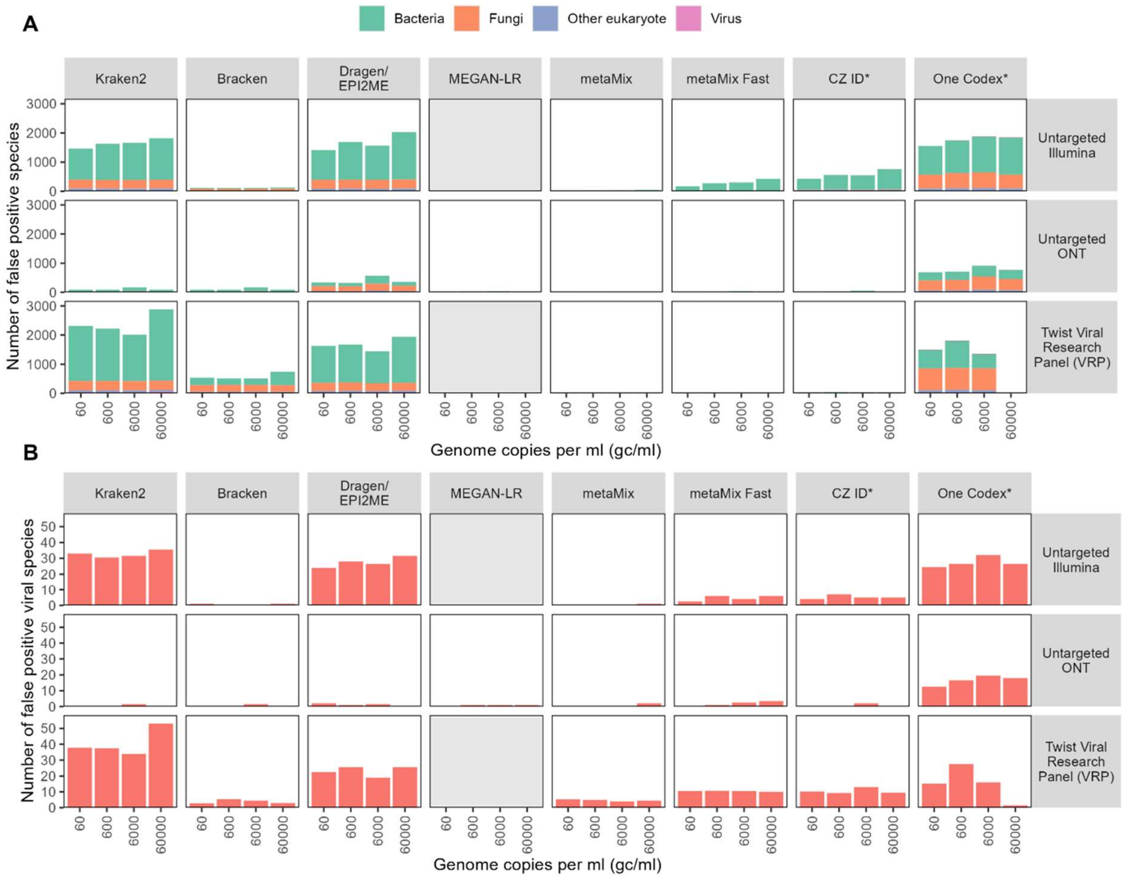


**SI Figure 1: False positive species without thresholds**

Number of false positive species, defined as a species that is classified as positive but not present in the mock community, for different taxonomic classifiers, by untargeted Illumina and ONT sequencing and capture probe enrichment with the Twist Biosciences Comprehensive Viral Research Panel followed by Illumina sequencing. **A** all species. **B** viruses only. Genome copy numbers refer to an average across the viral species – see **Supplementary Table 3**. Each bar shows the mean of at least two technical replicates.

Approaches used previously include raw read thresholds (2), proportion of total or microbial reads (2,5) and measures based on reads per million ratios (3). Whilst thresholds based on genome coverage may be informative, they are difficult to implement in an automated way for most of the classifiers tested here, which do not output genome alignments by default. Raw read thresholds, while simple to implement, may not be robust to differences in sequencing depth and host content, meaning that it may be preferable to use normalized measures such as reads per million (RPM). Comparison with the species in the negative control is important, but completely disregarding any species present at any in the negative control may be misleading, particularly if there have been low levels of cross-contamination between samples, which is common when viral loads are high. Reads per million ratios (RPMR) can be used to disregard species that are found at similar levels in the controls (3). Proportion of microbial reads (PMR) can be used to reject low level species that may be predicted due to misclassification, as well as being a more robust way of rejecting low-level species than raw read thresholds. Since we only had known true positives for viruses in this study, we used an RPMR of 10 and PMR of 0.01 for bacteria, fungi and other eukaryotes, which are thresholds that have been used in previous studies (2,4,6,7).

To determine the value of the thresholds for viruses, we first used all the data from all the nucleotide-based classifiers and sequencing runs (excluding the Twist VRP data with the standard One Codex database, which is not recommended and has low sensitivity when no thresholds are applied) tested to construct a receiver-operator characteristic (ROC) curve for RPMR, choosing the value where sensitivity was as close to specificity as possible (5.0, sensitivity = 86.0, specificity = 83.8%) (**SI Figure 2**). After implementing this threshold (accepting as positive any species that had no reads in the corresponding control) we then repeated this process to determine a threshold for PMR (0.00025). These thresholds resulted in a specificity and sensitivity of 87.4%. In our context, we would prefer to have better sensitivity, particularly as some false positive species are not clinically relevant or may be excluded using genome coverage thresholds. We therefore tested slightly decreased thresholds of RPMR = 5 and PMR = 0.0001, which provided a sensitivity of 91.7% and a specificity of 76.6% (**SI Figure 3**). We considered this to be a good balance between sensitivity and specificity for all classifiers except Dragen and One Codex for untargeted Illumina data. Use of RPMR = 5 and accepting any species that had no reads in the positive control gave a sensitivity of 99.5% but a specificity of only 28.6%, highlighting the benefit of using both measures. We also tested a logistic regression model to distinguish true positives. We used the same dataset to construct a model using RPMR and PMR. Setting the threshold to achieve a specificity of 91.7% resulted in a sensitivity of 77.4%, similar to use of combined RPMR and PMR thresholds as described above.

We also tested the published tool decontam (8) to help identify true positive species. Decontam uses the principle that contaminants are more abundant in negative controls (prevalence) and low concentration samples (frequency) (8). Since our negative controls consisted of human DNA and RNA, the input nucleic acid concentration was equal across all samples, meaning that frequency-based classification is not viable. We therefore used only the prevalence method, using the same input data as above, and produced a ROC curve of the threshold parameter. Sensitivity changed from 0 to 1 with the same specificity, 49.3% (**SI Figure 2**), making it slightly worse on our data than the method described above, which is to be expected since it is not designed to be used on our data type.

We therefore recommend using a combination of reads per million ratio and proportion of microbial reads to identify false positives for Illumina sequencing with tools that give a high number of false positives, such as Kraken2 and Illumina's Dragen Metagenomics pipeline. Exact thresholds used may vary between experimental setups and sample types but will usually need to be lower for viruses than for other organisms. ONT sequencing and Illumina sequencing with certain tools, such as metaMix, may be able to be used with only a simple comparison with the negative control, perhaps using RPMR alone.

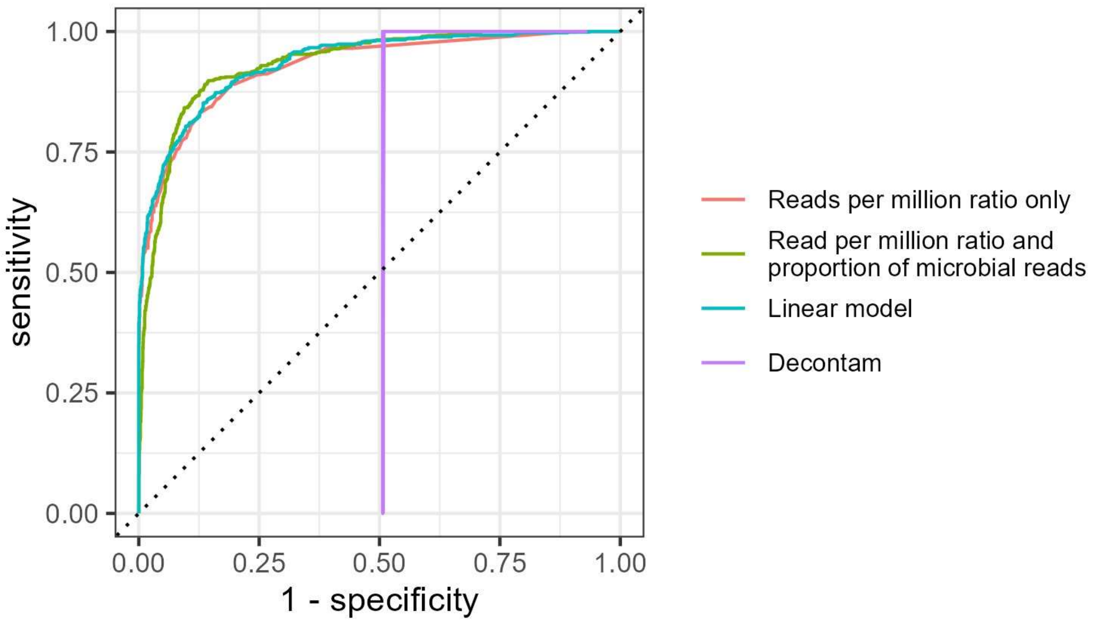


**SI Figure 2: Receiver operator characteristic curves**

ROC curves for the different sets of thresholds and models tested. The data used to generate Figure 1 was used for testing.

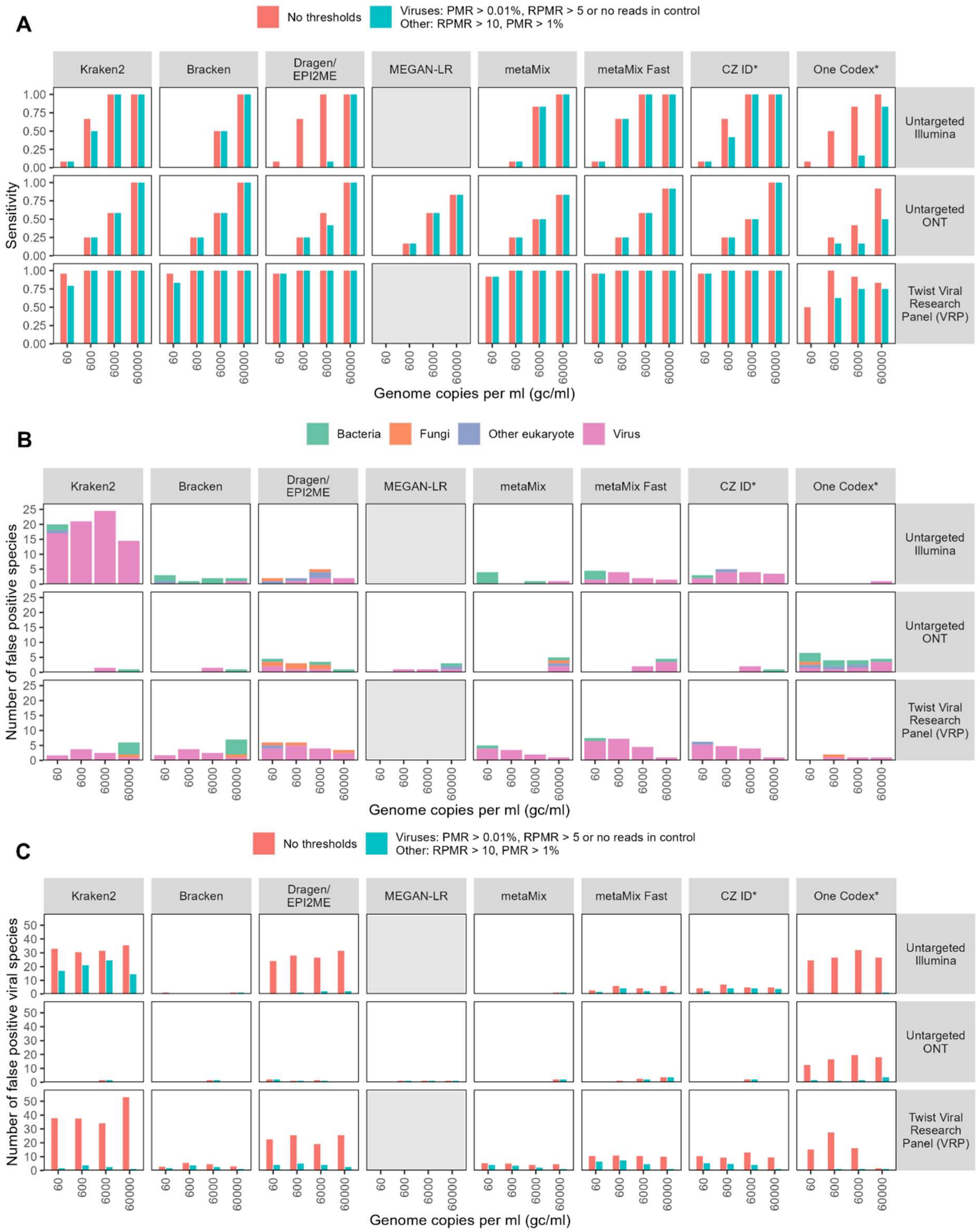


**SI Figure 3: Sensitivity and specificity with our suggested thresholds**

**A** Sensitivity to the species in the mock community before and after the application of thresholds, for different taxonomic classifiers, by untargeted Illumina and ONT sequencing and capture probe enrichment with the Twist Biosciences Comprehensive Viral Research Panel followed by Illumina sequencing. MEGAN-LR is only designed for ONT sequencing so was only run for this platform**. B,C** Number of false positive species, defined as a species that is classified as positive but not present in the mock community **B** False positive species from the raw output of the taxonomic classifiers with our thresholds applied. **C** Comparison of the numbers of viral positive species identified before and after the application of thresholds. Genome copy numbers refer to an average across the viral species – see **Supplementary Table 3**. Each bar shows the mean of at least two technical replicates**.**

**References**


1. Eisenhofer R, Minich JJ, Marotz C, Cooper A, Knight R, Weyrich LS. Contamination in Low Microbial Biomass Microbiome Studies: Issues and Recommendations. Trends in Microbiology. 2019 Feb 1;27(2):105–17.

2. Charalampous T, Alcolea-Medina A, Snell LB, Alder C, Tan M, Williams TGS, et al. Routine Metagenomics Service for ICU Patients with Respiratory Infection. Am J Respir Crit Care Med. 2024 Jan 15;209(2):164–74.

3. Gu W, Deng X, Lee M, Sucu YD, Arevalo S, Stryke D, et al. Rapid pathogen detection by metagenomic next-generation sequencing of infected body fluids. Nat Med. 2021 Jan;27(1):115–24.

4. Atkinson L, Lee JCD, Lennon A, Shah D, Storey N, Morfopoulou S, et al. Untargeted metagenomics protocol for the diagnosis of infection from CSF and tissue from sterile sites. Heliyon. 2023 Sep 1;9(9):e19854.

5. Portik DM, Brown CT, Pierce-Ward NT. Evaluation of taxonomic classification and profiling methods for long-read shotgun metagenomic sequencing datasets. BMC Bioinformatics. 2022 Dec 13;23(1):541.

6. Jia X, Hu L, Wu M, Ling Y, Wang W, Lu H, et al. A streamlined clinical metagenomic sequencing protocol for rapid pathogen identification. Sci Rep. 2021 Feb 23;11(1):4405.

7. Qin H, Peng J, Liu L, Wu J, Pan L, Huang X, et al. A Retrospective Paired Comparison Between Untargeted Next Generation Sequencing and Conventional Microbiology Tests With Wisely Chosen Metagenomic Sequencing Positive Criteria. Front Med (Lausanne). 2021 Oct 6;8:686247.

8. Davis NM, Proctor DM, Holmes SP, Relman DA, Callahan BJ. Simple statistical identification and removal of contaminant sequences in marker-gene and metagenomics data. Microbiome. 2018 Dec 17;6(1):226.
